# Decoding accelerometry for classification and prediction of critically ill patients with severe brain injury

**DOI:** 10.1101/2021.05.19.21257319

**Authors:** Shubhayu Bhattacharyay, John Rattray, Matthew Wang, Peter H. Dziedzic, Eusebia Calvillo, Han B. Kim, Eshan Joshi, Pawel Kudela, Ralph Etienne-Cummings, Robert D. Stevens

**Affiliations:** Laboratory of Computational Intensive Care Medicine, Johns Hopkins University, Baltimore, MD, USA; Department of Clinical Neurosciences, University of Cambridge, Cambridge, UK; Department of Biomedical Engineering, Johns Hopkins University, Baltimore, MD, USA; Department of Applied Mathematics and Statistics, Johns Hopkins University, Baltimore, MD, USA; Department of Electrical and Computer Engineering, Johns Hopkins University, Baltimore, MD, USA; Department of Anesthesiology and Critical Care Medicine, Johns Hopkins University, Baltimore, MD, USA; Department of Neurosurgery, Johns Hopkins University, Baltimore, MD, USA; Department of Neurology, Johns Hopkins University, Baltimore, MD, USA

## Abstract

Our goal is to explore quantitative motor features in critically ill patients with severe brain injury (SBI). We hypothesized that computational decoding of these features would yield information on underlying neurological states and outcomes. Using wearable microsensors placed on all extremities, we recorded a median 24.1 (IQR: 22.8–25.1) hours of high-frequency accelerometry data per patient from a prospective cohort (*n* = 69) admitted to the ICU with SBI. Models were trained using time-, frequency-, and wavelet-domain features and levels of responsiveness and outcome as labels. The two primary tasks were detection of levels of responsiveness, assessed by motor sub-score of the Glasgow Coma Scale (GCSm), and prediction of functional outcome at discharge, measured with the Glasgow Outcome Scale–Extended (GOSE). Detection models achieved significant (AUC: 0.70 [95% CI: 0.53–0.85]) and consistent (observation windows: 12 min – 9 hours) discrimination of SBI patients capable of purposeful movement (GCSm > 4). Prediction models accurately discriminated patients of upper moderate disability or better (GOSE > 5) with 2–6 hours of observation (AUC: 0.82 [95% CI: 0.75–0.90]). Results suggest that time series analysis of motor activity yields clinically relevant insights on underlying functional states and short-term outcomes in patients with SBI.

## INTRODUCTION

Severe brain injury (SBI), defined as an acute injury to or illness in the brain that impairs consciousness, imposes the greatest global burden of mortality, long-term disability, and economic cost among all major injury types [1]. Despite other advances in intensive care medicine, existing approaches to predict SBI outcomes, such as recovery of consciousness and functional independence, in the intensive care unit (ICU) are imprecise for individual patients [2] and can raise ethical concerns due to the potential for withdrawal of life-sustaining therapies (WLST) [3]. For example, both general ICU outcome prediction models – e.g., the Acute Physiologic Assessment and Chronic Health Evaluation [4] (APACHE) II – and models developed for specific types of SBI – e.g., the International Mission for Prognosis and Analysis of Clinical Trials in Traumatic Brain Injury [5] (IMPACT) model – are calculated at 24 hours after ICU admission and thus disregard the dynamic, heterogenous pathophysiological process that unfolds after SBI [6,7]. At the same time, recent developments in artificial intelligence and big data processing represent an opportunity to enhance SBI patient monitoring with high-resolution, longitudinal waveform data and to improve the precision of SBI prognostication with flexible modeling strategies [8]. Hence, a key focus in the care of SBI is the discovery and validation of quantitative monitoring modalities that improve upon the precision of clinical characterization and the accuracy and reliability of predicted outcomes [9].

For acute neurological disorders, the assessment of motor function provides a unique window into neural systems associated with sensorimotor processing, emotion, coordination, planning, and learning [10–12]. Neurological damage and ICU practices (e.g., sedation, bedrest) are associated with a dramatic reduction in normal physical activity [13], which may result in systemic pro-inflammatory signaling [14] and an elevated risk of venous thromboembolism, infection, skin and soft tissue damage, delirium, and loss of muscle mass and strength [15–18]. A corollary is that structured programs designed to increase physical activity for SBI patients in the ICU can significantly reduce neurological complications and may lead to improved functional recovery [19]. However, it is uncertain whether the computational analysis of continuously recorded motion in the ICU could yield clinically significant gains for SBI monitoring and prognosis.

Wearable accelerometers provide an objective and continuous assessment of motor activity over extended periods of time [20]. In contrast to most other motion sensing modalities [21], integration of accelerometers in the ICU is feasible. Advances in microelectromechanical systems (MEMS) technology have made it possible to construct inexpensive, minimally obtrusive wearable accelerometers that can be optimized for the clinical space [22]. Accelerometers respond to changes in movement frequency and intensity, measure tilt from the gravitational axis, and produce little variation or drift over time [22–26]. The use of accelerometers to monitor gross physical activity in the ICU has already been tested with varying degrees of success [27]. Herein, we aim to more specifically determine whether a relationship exists between motion features derived from triaxial accelerometry time-series and neurological motor states and functional outcomes of SBI patients.

In this pilot study of the Neurological Injury Motion Sensing (NIMS) project, we explore the impact and limitations of high-resolution accelerometry in patients with SBI admitted to the ICU. We developed a matrix of wearable accelerometers to quantitatively capture motor activity from the extremities of SBI patients. Applying techniques from time-series analysis, dimensionality reduction, and logistic regression, we extract interpretable time-, frequency-, and wavelet-domain motion features and assess their performance in motor function detection and short- and long-term functional outcome prediction models. We then assess relative significance of the extracted features to determine how specific accelerometry profiles relate to clinically evaluated motor function and global outcomes. Finally, through a retrospective case analysis, we demonstrate how accelerometry-based model outputs can potentially be used to monitor neurological transitions.

## RESULTS

### Study population characteristics

Of the 72 total SBI patients recruited in the ICU, 3 participants were excluded from the study due to withdrawn consent (*n* = 2) or corruption of accelerometry data during upload (*n* = 1), resulting in a study population of *n* = 69. Five patients were lost from one-year follow-up due to unsuccessful contact, and thus the study population at 12 months post hospital discharge was *n* = 64. A Consolidated Standards of Reporting Trials [28] (CONSORT)-style flow diagram for patient enrollment and follow-up is provided in **Supplementary Figure S1 online**, and detailed characteristics of the study population are summarized in **Table 1**.

**Table 1.** Study population characteristics.

| Characteristic |  | Severe brain injury patients (*n* = 69) |
| --- | --- | --- |
| Age (y) |  | 59 (48–70) |
| M/F (*n*) |  | 33/36 |
| Types of severe brain injury (*n*) | Intracranial hemorrhage (ICH) | 29 (42.03%) |
|  | Subdural or epidural hematoma (SDH/EDH) | 18 (26.09%) |
|  | Subarachnoid hemorrhage (SAH) | 17 (24.64%) |
|  | Cerebrovascular accident (CVA) | 14 (20.29%) |
|  | Brain tumor (BT) | 12 (17.39%) |
|  | Traumatic brain injury (TBI) | 8 (11.59%) |
| Motor component score of the Glasgow Coma Scale (GCSm) at ICU admission | (1) No response | 4 (5.80%) |
|  | (2) Abnormal extension | 3 (4.35%) |
|  | (3) Abnormal flexion | 6 (8.70%) |
|  | (4) Withdrawal from stimulus | 4 (5.80%) |
|  | (5) Movement localized to stimulus | 17 (24.64%) |
|  | (6) Obeys commands | 35 (50.72%) |
| Motor component score of the Glasgow Coma Scale (GCSm) at ICU discharge | (1) No response | 7 (10.14%) |
|  | (2) Abnormal extension | 5 (7.25%) |
|  | (3) Abnormal flexion | 4 (5.80%) |
|  | (4) Withdrawal from stimulus | 4 (5.80%) |
|  | (5) Movement localized to stimulus | 11 (15.94%) |
|  | (6) Obeys commands | 38 (55.07%) |
| Net change in GCSm during ICU stay |  | 0 (0–+1) |
| Glasgow Outcome Scale - Extended (GOSE) at hospital discharge | (1) Dead | 16 (23.19%) |
|  | (2) Vegetative state | 4 (5.80%) |
|  | (3) Lower severe disability | 30 (43.48%) |
|  | (4) Upper severe disability | 11 (15.94%) |
|  | (5) Lower moderate disability | 6 (8.70%) |
|  | (6) Upper moderate disability | 1 (1.45%) |
|  | (7) Lower good recovery | 1 (1.45%) |
|  | (8) Upper good recovery | 0 (0%) |
| Glasgow Outcome Scale - Extended (GOSE) at 12 months post discharge* | (1) Dead | 28 (43.75%) |
|  | (2) Vegetative state | 2 (3.12%) |
|  | (3) Lower severe disability | 14 (21.88%) |
|  | (4) Upper severe disability | 12 (18.75%) |
|  | (5) Lower moderate disability | 3 (4.69%) |
|  | (6) Upper moderate disability | 0 (0%) |
|  | (7) Lower good recovery | 3 (4.69%) |
|  | (8) Upper good recovery | 2 (3.12%) |
| Net change in GOSE in 12 months post discharge* |  | 0 (–3.425–+3) |
| Acute Physiology and Chronic Health Evaluation (APACHE) II at 24 hours post ICU admission | Score (0–71) | 21 (16–25) |
|  | Predicted risk of in-hospital mortality (%) | 46.00 (29.30–67.00) |
|  | Accuracy of in-hospital mortality prediction | 0.70 |
|  | AUC<sup>†</sup> of in-hospital mortality prediction | 0.84 |
| Length of stay in ICU (d) |  | 19 (11–29) |
| Delay between ICU discharge and recording start (d) |  | 7 (2–15) |
| Recording duration (h) |  | 24.09 (22.81–25.11) |
| Percentage of ICU stay recorded (%) |  | 5.54 (3.13–8.51) |
| Percentage of ICU stay elapsed before recording start (%) |  | 43.53 (24.98–62.88) |
| Recording duration (h) |  | 24.09 (22.81–25.11) |

Values represent medians with interquartile ranges (Q1–Q3) in parentheses or counts with percentages (%) in parentheses.
\*Total sample size at 12 months post discharge is *n* = 64.
<sup>†</sup>Area under the receiver operating characteristic curve, i.e., the probability that the predicted mortality risk of a randomly chosen patient who died is greater than the predicted mortality risk of a randomly chosen patient who survived hospital stay.

From each of the study participants, we collected triaxial accelerometry data (sampled at 10 Hz) from a wearable matrix of 6 sensors, placed on each elbow, wrist, and ankle and an additional sensor placed on the bed for external movement correction (**Fig. 1a**). The median recording duration per patient was 24.09 hours (IQR: 22.81–25.11 hours), and accelerometry data was recorded fairly uniformly across the stages of ICU stay in terms of proportion completed (**Supplementary Fig. S2 online**). In total, 1,701 hours of multisegmental accelerometry data were recorded.

**Fig. 1.**
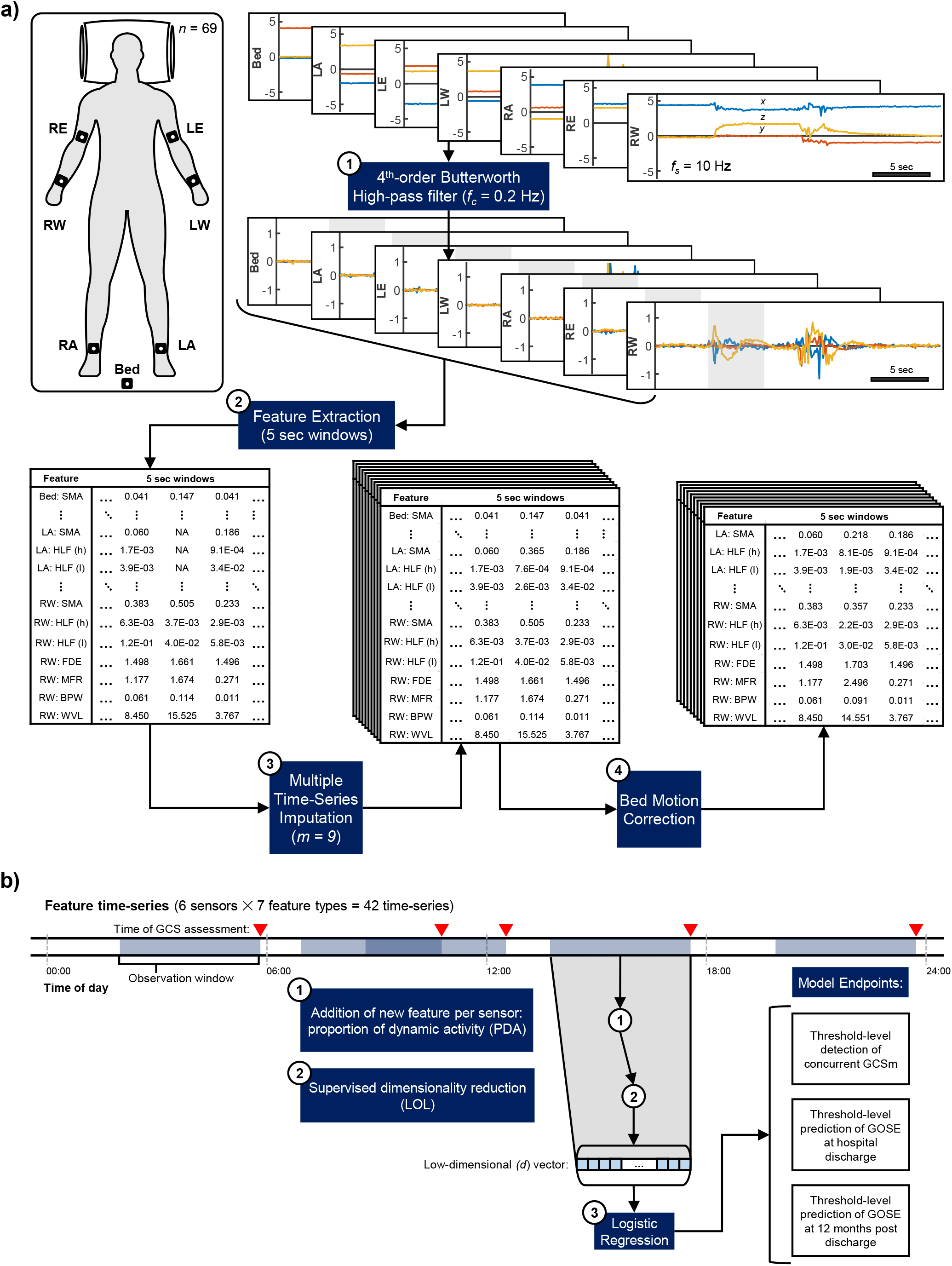
Accelerometry processing and feature extraction pipeline and experimental paradigm. **a** Accelerometry (top right, units: *g*) was continuously captured from wearable sensors placed on six joints of severe brain injury patients (*n* = 69) in the ICU (top left) for a median of 24 (IQR: 23 - 25) hours per patient. Sensor placement acronyms correspond to the right and left elbows (RE and LE), the right and left wrists (RW and LW), and the right and left ankles (RA and LE). *f*_*s*_ represents the sampling rate of accelerometry in Hz. The raw accelerometry collected from each patient underwent a four-step (numbered boxes) preprocessing pipeline before being transformed into a complete, multiply imputed (*m* = 9) feature set for analysis. Feature type acronyms are decoded in **Table 3**, and the steps of the processing and extraction pipeline are described in the **Methods** section. **b** Experimental paradigm to derive model probabilities for motor function detection per the motor component score of the Glasgow Coma Scale (GCSm) and functional outcome predictions per the Glasgow Outcome Scale – Extended (GOSE). GCSm evaluations were reported in the patients’ electronic health records by ICU clinicians and may have occurred at any time during ICU stay (red, upside-down triangle). We tested 19 distinct observation windows (light-blue, shaded regions), ranging from 3 minutes to 24 hours (**Supplementary Table S1 online**). The motion feature time-series (end of pipeline in **a**) in the observation window preceding each GCSm evaluation underwent two more processing steps: (1) the calculation and addition of another feature representing the proportion of dynamic activity (PDA) of each sensor in the observation window, and (2) supervised dimensionality reduction, in which a linear optimal low-rank projection (LOL) matrix is learned from the training set to exploit the variance in the dataset, stratified by model endpoint, and output the best-discriminating low (*d*, from 2 to 20) dimensional vector (see **Methods**). These vectors were then used to (3) train logistic regression models that, on a threshold-level, detected the concurrent GCSm or predicted GOSE at hospital discharge or at 12 months post discharge.

During their stay in the ICU (median: 19 days, IQR: 11–29 days), study participants were evaluated with the Glasgow Coma Scale (GCS) [29,30] a median 9.25 times per day (IQR: 7.17–11.50 times per day). In total, we extracted scores from 14,240 GCS evaluations, 13,190 of which (92.63%) took place in the ICU and 653 of which (4.59%) coincided with accelerometry capture times. The individual trajectory of the motor component scores of the GCS (GCSm), along with corresponding times of accelerometry capture, of each patient included in our analysis is provided in **Supplementary Figure S3 online**.

### Motor function detection performance

In this work, we use clinically evaluated GCSm scores extracted from electronic health records (EHR) at any point during ICU stay as the primary labels of functional motor states. The scores of the 6-point GCSm are defined by best motor responses obtained spontaneously and to graded physical stimuli and are outlined in **Table 1**.

We trained and evaluated threshold-level GCSm detection models from automated accelerometry-based motion features extracted from 19 varying observation windows, from 3 minutes to 24 hours, directly preceding the GCSm evaluations (**Fig. 1b**). The count distributions of GCSm scores available for each observation window are listed in **Supplementary Table S1 online**.

The receiver operating characteristic (ROC) curves of the optimally discriminating models at each GCSm threshold, along with their mean areas under the curves (AUC) and optimal observation windows, are shown in **Figure 2a**. Based on the 95% confidence intervals of mean AUC, significant discrimination (AUC > 0.5, α = 0.05) was achieved by the extracted features at every threshold of GCSm except for GCSm > 2. However, only GCSm > 4 detection models achieve significant discrimination from shorter observation window durations (≤ 30 minutes); GCSm > 4 detection models achieve significant discrimination consistently with an observation window of 12 minutes or greater (**Fig. 2b**). The mean AUCs, along with 95% confidence intervals, at each threshold of GCSm is provided for all 19 tested observation windows in **Supplementary Table S2 online**. As GCSm > 1, GCSm > 3, and GCSm > 5 detection models achieve significant discrimination at less than or equal to 3 different observation windows, only GCSm > 4 detection models achieve significant discrimination at a broad range of observation windows (12 min – 9 hours). Binary classification performance metrics of optimally discriminating motor function detection models are provided in **Table 2**. At none of the GCSm thresholds do the models achieve significantly greater accuracy than the proportion of the most represented class based on 95% confidence intervals. Only the GCSm > 4 detection model achieves a higher mean accuracy (0.71) and a significantly greater F_1_ score (0.78 [95% CI: 0.67–0.87]) than its proportion of the most represented (in this case, positive) class (0.66). Only GCSm > 4 and GCSm > 5 detection models achieved both a mean sensitivity and mean specificity over 0.5, but not significantly.

**Fig. 2.**
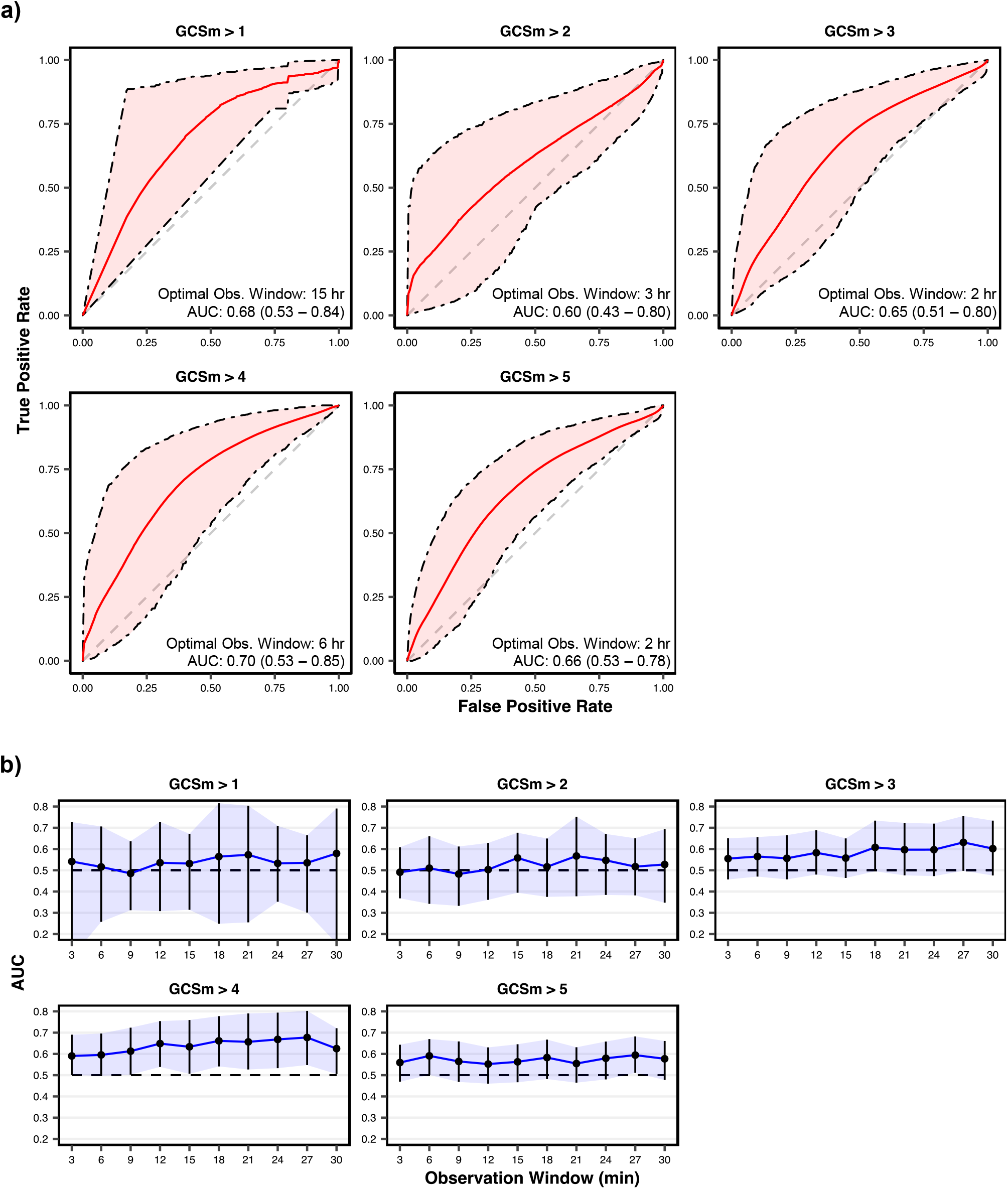
Discrimination performance of motor function detection models on validation sets. **a** Receiver operating characteristic (ROC) curves of models pertaining to the observation windows with the highest achieved area under the ROC curve (AUC) per each detection threshold of the motor component score of the Glasgow Coma Scale (GCSm). AUC corresponds to the probability that the model can correctly discriminate a randomly selected patient above the threshold from a randomly selected patient below the threshold. Shaded areas are 95% confidence intervals derived using bias-corrected bootstrapping (1,000 resamples) to represent the variation across repeated cross-validation folds (5 repeats of 5 folds) and nine missing value imputations. The values in each box represent the observation window achieving the highest AUC as well as the corresponding mean AUC (with 95% confidence interval in parentheses). The diagonal dashed line represents the line of no discrimination (AUC = 0.5). **b** AUC vs. observation windows up to 30 minutes per each detection threshold of the motor component score of the Glasgow Coma Scale (GCSm). Points represent observation windows tested and error bars (with the associated shaded region) represent the 95% confidence interval. The horizontal dashed line corresponds to no discrimination (AUC = 0.5).

**Table 2.**
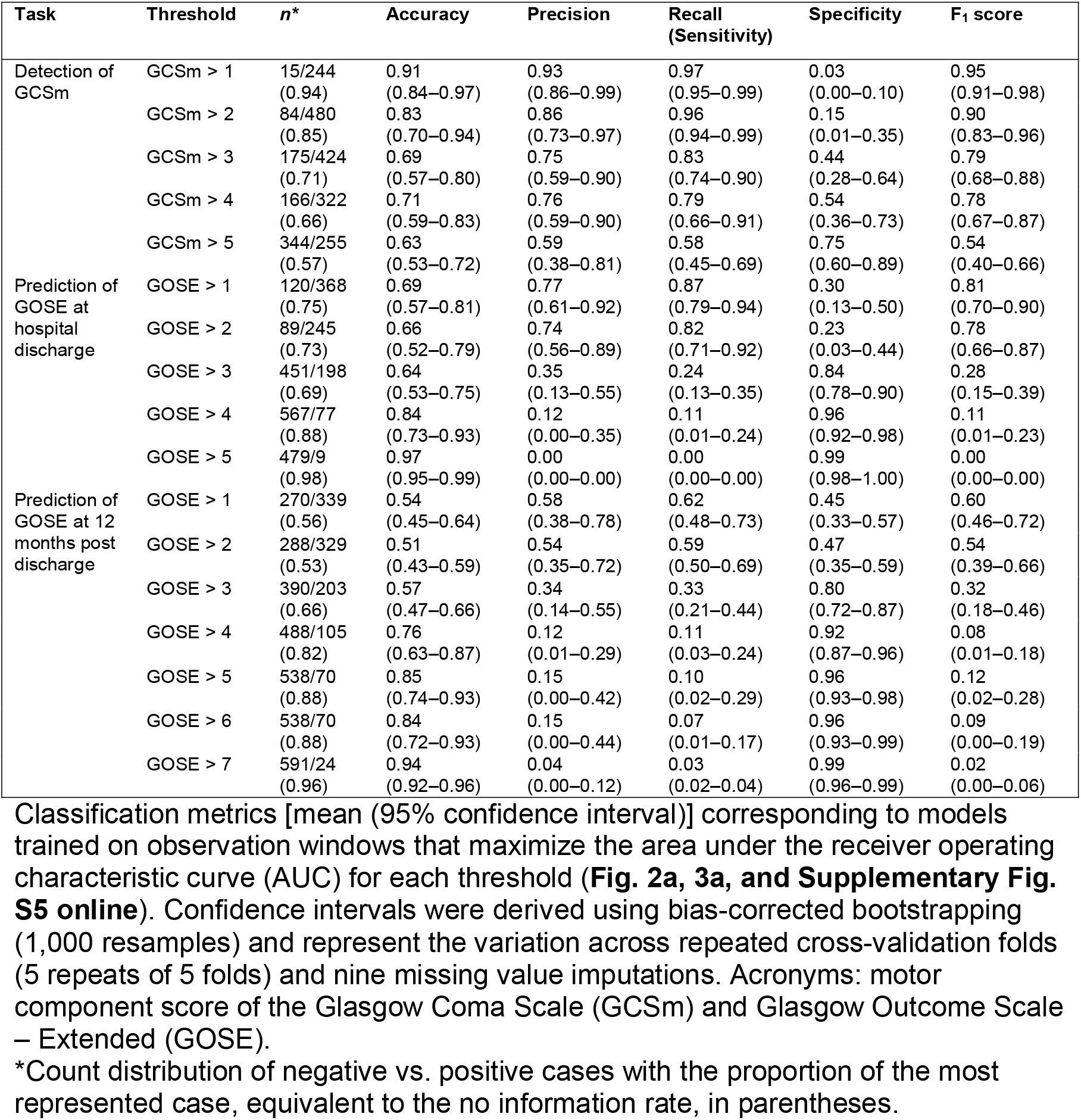
Classification performance metrics of optimally discriminating models.

### Functional outcome at hospital discharge prediction performance

We used clinically evaluated Glasgow Outcome Scale – Extended (GOSE) scores as the primary indicator of functional outcomes, both at hospital discharge and at 12 months post discharge. The scores of the 8-point GOSE are outlined in **Table 1**.

We trained and evaluated threshold-level GOSE at hospital discharge prediction models from automated accelerometry-based motion features extracted from the same 19 varying observation windows directly preceding GCSm evaluations (**Fig. 1b**). The median lead window duration (i.e., time between end of observation window and hospital discharge) was 20 days (IQR: 10–33 days). The count distributions of GOSE scores, at discharge, available for each observation window are listed in **Supplementary Table S3 online**. Given the low proportion of patients (1.45%) with good recovery (GOSE > 6) at hospital discharge, we limited our threshold-level analysis to GOSE > 1, GOSE > 2, GOSE > 3, GOSE > 4, and GOSE > 5.

The receiver operating characteristic (ROC) curves of the optimally discriminating models at each GOSE threshold, along with their mean areas under the curves (AUC) and optimal observation windows, are shown in **Figure 3a**. Based on the 95% confidence intervals of mean AUC, significant discrimination (AUC > 0.5, α = 0.05) was achieved by the extracted features only at GOSE > 5. GOSE > 5 prediction models achieve significant discrimination at observation windows of two hours or greater, with a peak mean AUC of 0.82 (95% CI: 0.75–0.90) at an observation window duration of 6 hours (**Fig. 3b**). The mean AUCs, along with 95% confidence intervals, at each tested threshold of GOSE is provided for all 19 tested observation windows in **Supplementary Table S4 online**.

**Fig. 3.**
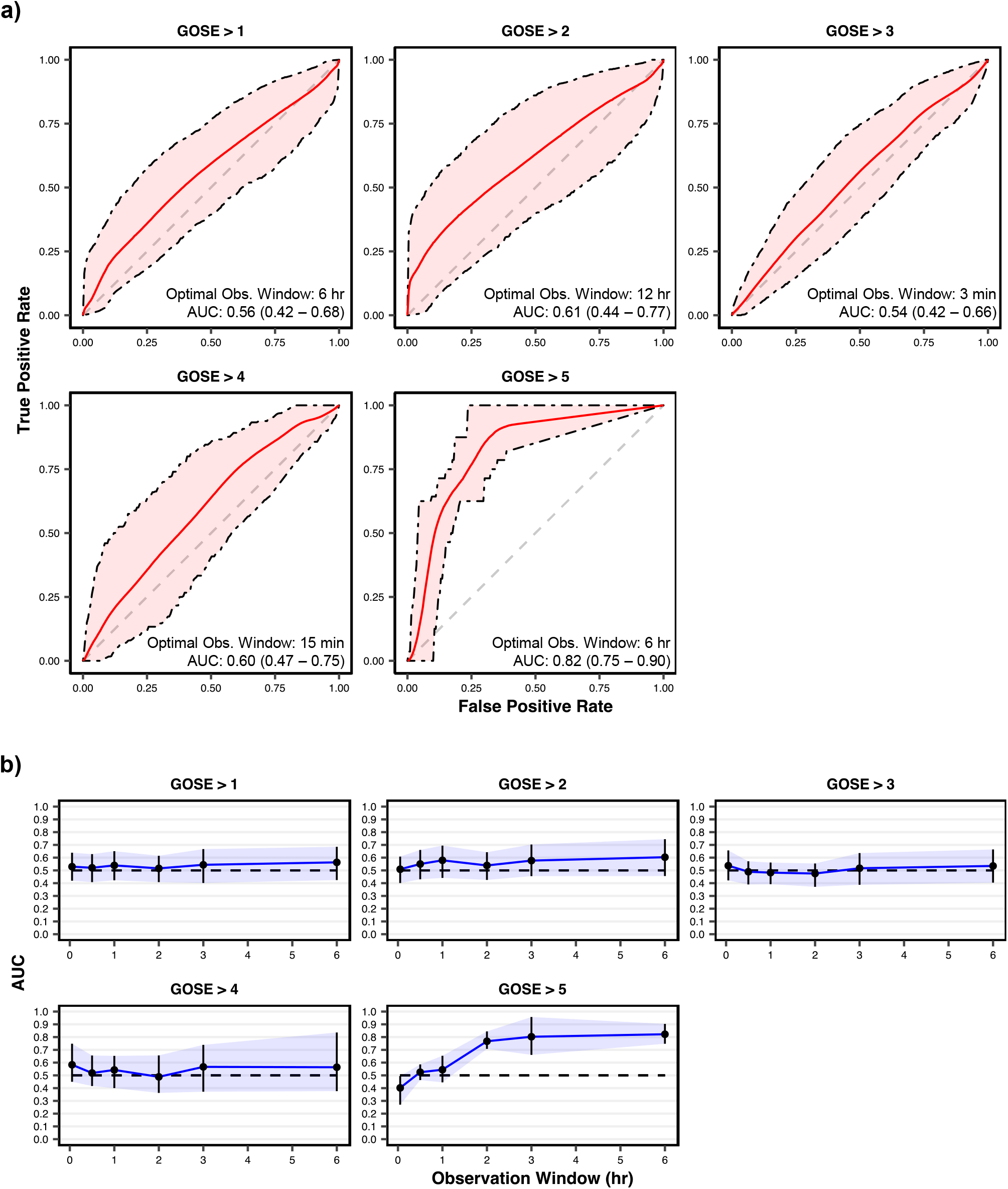
Discrimination performance of functional outcome at hospital discharge prediction models on validation sets. **a** Receiver operating characteristic (ROC) curves of models pertaining to the observation windows with the highest achieved area under the ROC curve (AUC) per each tested prediction threshold of the Glasgow Outcome Scale – Extended (GOSE). AUC corresponds to the probability that the model can correctly discriminate a randomly selected patient above the threshold from a randomly selected patient below the threshold. Shaded areas are 95% confidence intervals derived using bias-corrected bootstrapping (1,000 resamples) to represent the variation across repeated cross-validation folds (5 repeats of 5 folds) and nine missing value imputations. The values in each box represent the observation window achieving the highest AUC as well as the corresponding mean AUC (with 95% confidence interval in parentheses). The diagonal dashed line represents the line of no discrimination (AUC = 0.5). **b** AUC vs. observation windows up to 6 hours per each tested prediction threshold of the Glasgow Outcome Scale – Extended (GOSE). Points represent observation windows tested and error bars (with the associated shaded region) represent the 95% confidence interval. The horizontal dashed line corresponds to no discrimination (AUC = 0.5).

Binary classification performance metrics of optimally discriminating functional outcome prediction models are provided in **Table 2**. At none of the GOSE thresholds do the models achieve a significantly greater F_1_ score than the proportion of the positive class or a greater mean accuracy than the proportion of the most represented class. Despite its strong discrimination performance, the GOSE > 5 prediction model had low precision and sensitivity. From the precision recall curve for this model (**Supplementary Fig. S4 online**), we observe a mean average precision of 0.08 (95% CI: 0.02–0.18), which, while low, is significantly greater than the proportion of the positive class (0.02). This indicates, that while prediction probabilities for true positive cases are, on average, greater than prediction probabilities for true negative cases, they seldom cross the 0.5 threshold for proper classification (**Supplementary Fig. S4 online**).

### Functional outcome at 12 months post discharge prediction performance

We trained and evaluated threshold-level GOSE at 12 (±1) months post hospital discharge prediction models from automated accelerometry-based motion features extracted from the same 19 varying observation windows directly preceding GCSm evaluations (**Fig. 1b**). The count distributions of GOSE scores, at 12 months, available for each observation window are listed in **Supplementary Table S5 online**. The receiver operating characteristic (ROC) curves of the optimally discriminating models at each GOSE threshold, along with their mean areas under the curves (AUC) and optimal observation windows, are shown in **Supplementary Figure S5 online**. Based on the 95% confidence intervals of mean AUC, significant discrimination (AUC > 0.5, α = 0.05) was not achieved by the extracted features at any of the GOSE thresholds. Mean AUC is largely independent of observation window duration at each of the thresholds (**Supplementary Fig. S5 online**). The mean AUCs, along with 95% confidence intervals, at each threshold of GOSE is provided for all 19 tested observation windows in **Supplementary Table S6 online**. Binary classification performance metrics of optimally discriminating functional outcome prediction, at 12 months post discharge, models are provided in **Table 2**.

### Calibration of motor function detection and functional outcome prediction

The probability calibration curves and associated prediction distributions of the optimally discriminating models at each threshold for GCSm detection and GOSE (at hospital discharge) prediction are provided in **Supplementary Figure S6 online**. We observe that the GCSm > 4 detection model achieves the best graphical model calibration of all those tested (*E*_*max*_ = 0.30 [95% CI: 0.08–0.64]). However, when considering the prevalence of predicted probabilities in calibration assessment with the integrated calibration index (ICI) [31], we observe that the GOSE > 5 prediction model has the most ideal calibration (ICI = 0.01 [95% CI: 0.00–0.02]). The discrepancy between the weighted and graphical calibration of GOSE > 5 indicates a strong class imbalance, suggesting that more positive cases are necessary to train and recalibrate this model for proper classification. Probability calibration metrics of all optimally discriminating models are provided in **Supplementary Table S7 online**.

### Extracted feature and sensor placement analysis

At the end of our accelerometry processing pipeline (**Fig. 1**), we extracted eight unique feature types (**Table 3**) from each of the six accelerometers placed on SBI patient joints. For each of these 48 feature-sensor combinations, we calculate a relative significance score equivalent to the mean absolute value of the learned coefficients of supervised dimensionality reduction (i.e., the relative importance in explaining the variance in the dataset stratified by the endpoint) weighted by the absolute value of learned logistic regression coefficients (see **Methods**).

**Table 3.** Overview of extracted motion feature types.

| Acronym | Feature description | Domain | Reference |
| --- | --- | --- | --- |
| PDA | Proportion of dynamic activity ($SMA \geq 0.135g$) in observation window | Time | [32] |
| SMA | Signal magnitude area | Time | [59] |
| HLF (h) | Median of high-pass-filtered (4th-order Butterworth, $f_c = 2.5$ Hz) signal | Time | [60] |
| HLF (l) | Median of low-pass-filtered (4th-order Butterworth, $f_c = 2.5$ Hz) signal | Time | |
| MFR | Median frequency according to Fourier transform coefficients | Frequency | [61] |
| FDE | Frequency-domain entropy | Frequency | [62] |
| BPW | Band power between 0.3 and 3.5 Hz | Frequency | [63] |
| WVL | Level 2 – 6 detail coefficients of the 5th-order Daubechies wavelet transform | Wavelets | [64] |

Each feature, except PDA, was extracted from non-overlapping 5 second windows and root-sum-of-squares leveled across the three axes of accelerometry measurement (x, y, z). A single PDA value was calculated per each sensor for the entire observation window. $f_c$ represents the critical frequency of the Butterworth filter. The references point to original works in which the features (or similar variants) were used in physical activity recognition. Explicit formulae for each feature can be found in **Methods**.

We consider the optimally discriminating configurations of the two most promising model types as representatives for motor function detection and functional outcome prediction respectively: (a) GCSm > 4 with a 6-hour observation window and (b) GOSE (at hospital discharge) > 5 with a 6-hour observation window. The feature significance scores of these two model types are visualized as heatmaps in **Fig. 4a** and **Fig. 4b** respectively.

**Fig. 4.**
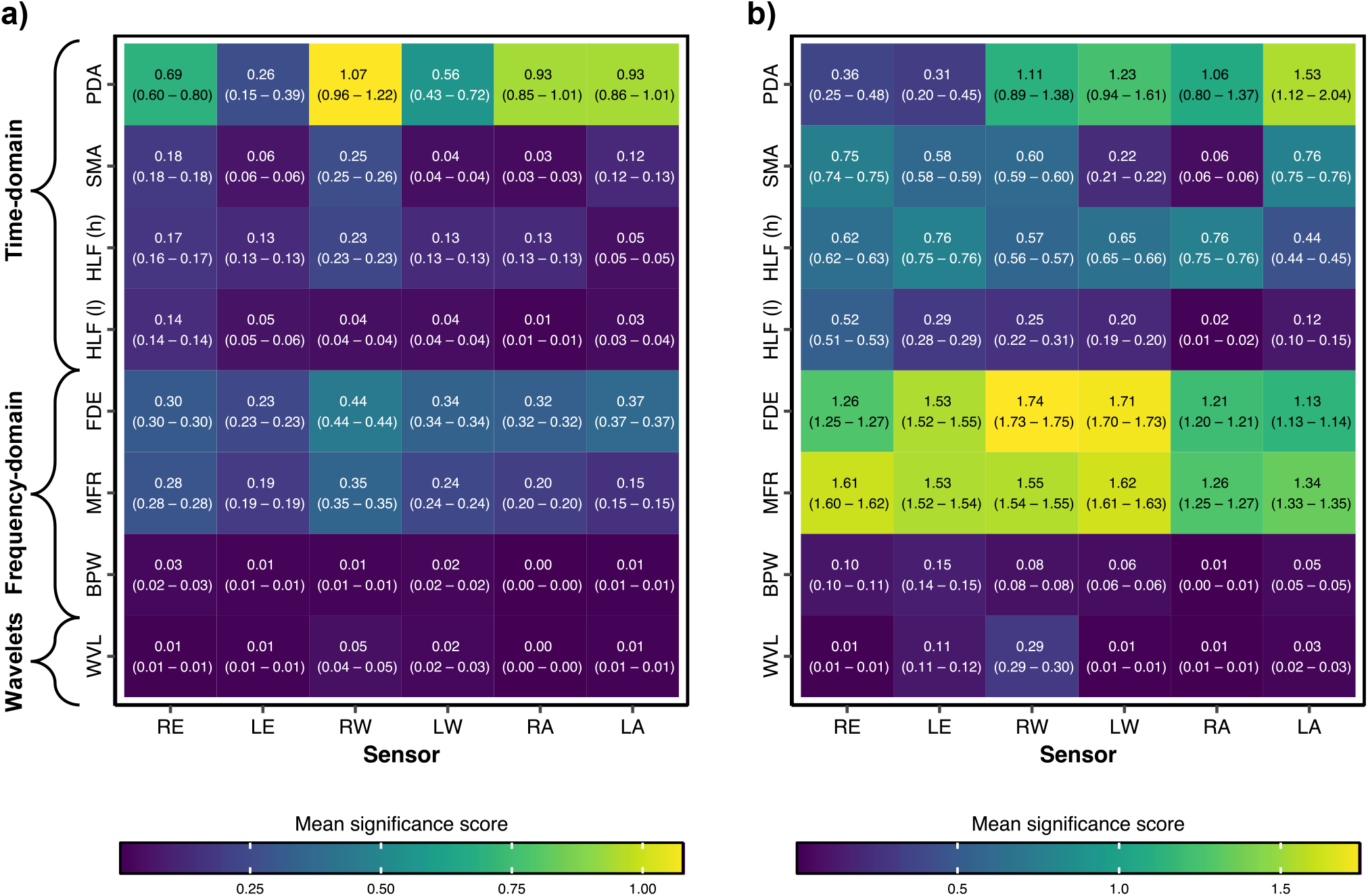
Feature significance matrices of optimally discriminating motor function detection and functional outcome prediction models. Significance scores are calculated by weighting linear optimal low-rank projection (LOL) coefficients of sensor-feature type combinations – which represent the relative importance of each timestep of each sensor-feature type combination in explaining the variance in the dataset stratified by (**a**) the motor component score of the Glasgow Coma Scale (GCSm) or (**b**) the Glasgow Outcome Scale – Extended (GOSE) – by the logistic regression coefficients of the corresponding LOL component in the low-dimensional vector (**Fig. 1b**). The higher (yellow) the mean significance score, the greater the combination of that sensor-feature type combination in the learned discrimination of patients at that threshold. The feature significance matrix in (**a**) corresponds to the optimally discriminating model configuration (6-hour observation window) for detection of GCSm > 4 (**Fig. 2a**) while the matrix in (**b**) corresponds to the optimally discriminating model configuration (6-hour observation window) for prediction of GOSE > 5 at hospital discharge (**Fig. 3a**). Mean significance scores (across all timesteps for a sensor-feature type combination) are listed as well as 95% confidence intervals bootstrapped from 1,000 resamples to represent the variation across repeated cross-validation folds (5 repeats of 5 folds) and nine missing value imputations. Sensor placement acronyms correspond to joints shown in **Fig. 1** and feature type acronyms are decoded in **Table 3**.

For both motor function detection and functional outcome prediction, there is more variation in significance scores across feature types than across sensor placements. For motor function detection, the proportion of dynamic activity (PDA) in the observation window, the frequency-domain entropy (FDE), and the median frequency (MFR) are the three most significant feature types, descending in that order. For functional outcome prediction, the descending order of the three most significant feature types is FDE, MFR, and PDA. PDA is a crude measurement of overall physical activity [32], while FDE enables differentiation between activity profiles which have simple acceleration patterns and those with more complex patterns [20]. From the pair of high-pass-filtered medians (HLF (h)) and low-pass-filtered medians (HLF (l)), HLF (h) has a significantly greater mean significance score than HLF (l) for every sensor placement in both model endpoints based on 95% confidence intervals. This, along with the relative significance of MFR, suggests that finer movements, captured in higher frequencies of accelerometry, can be more clinically significant in discriminating functional motor states and global outcomes from SBI. Moreover, the consistently strong significance of PDA, FDE, and MFR suggests that features of both the time domain (PDA) and the frequency domain (FDE, MFR) in combination may be useful for clinical assessments of functional neurological states.

In detecting motor function, the right wrist (RW) sensor was the most significant placement across the five most significant feature types. The trajectories of mean motion feature values in the six hours preceding GCSm evaluations (**Supplementary Fig. S7 online**) visually demonstrate that features extracted from the wrist-placed sensors better discriminate cases of GCSm 5 and 6 from the rest of the GCSm scores. This follows clinical observations of a greater frequency of conscious movement in hands and wrists of bedridden SBI patients during ICU stay. Moreover, abnormal profiles of flexion and extension, associated with SBI, are most often observed in the wrists, and thus, the wrist-placed sensors may be more sensitive to abnormal patterns of movement, corresponding to lower levels of consciousness, than the elbow- or ankle-placed sensors.

In functional outcome prediction, we observe the greatest significance scores ascribed to wrist-placed sensors (RW and LW) in the most significant frequency-domain features (FDE and MFR), but the ankle- (RA and LA) and elbow-placed sensors have the greatest significance scores in the most significant time-domain features (PDA, SMA, and HLF (h)). Wrist movements are finer than elbow and ankle movements and may be best distinguished in the frequency-domain in relation to global outcomes.

The correlation of each of the extracted motion features across the six sensor placements is visualized in **Supplementary Figure S8 online**, and violin plots of the distributions of motion features, stratified by GCSm, are presented in **Supplementary Figure S9 online**.

### Retrospective case study analysis of motor function detection in practice

The final goal of this study was to determine how the motion feature-based predictions would react, in real-time, to instances in which a patient’s functional motor state changed. From six of our study participants, we happened to have recorded accelerometry while they experienced at least one transition between GCSm > 4 and GCSm ≤ 4 with at least six hours of recording (for an observation window) before the transition(s). For each of these patients, we trained two different GCSm > 4 detection models on the remaining patient set: one with the best short (<30 minutes) observation window (27 minutes) and one with the best long (≥1 hour) observation window (6 hours) based on overall discrimination performance (**Supplementary Table S2**). We selected two distinct observation windows to understand the effect of window duration on responsiveness to motor state transition. With all of the trained models, we returned predictions with a sliding window (step: 10 minutes) on the corresponding case study patient to retrospectively examine the trajectories of detection probabilities against the recorded times of GCSm transition (**Fig. 5**).

**Fig. 5.**
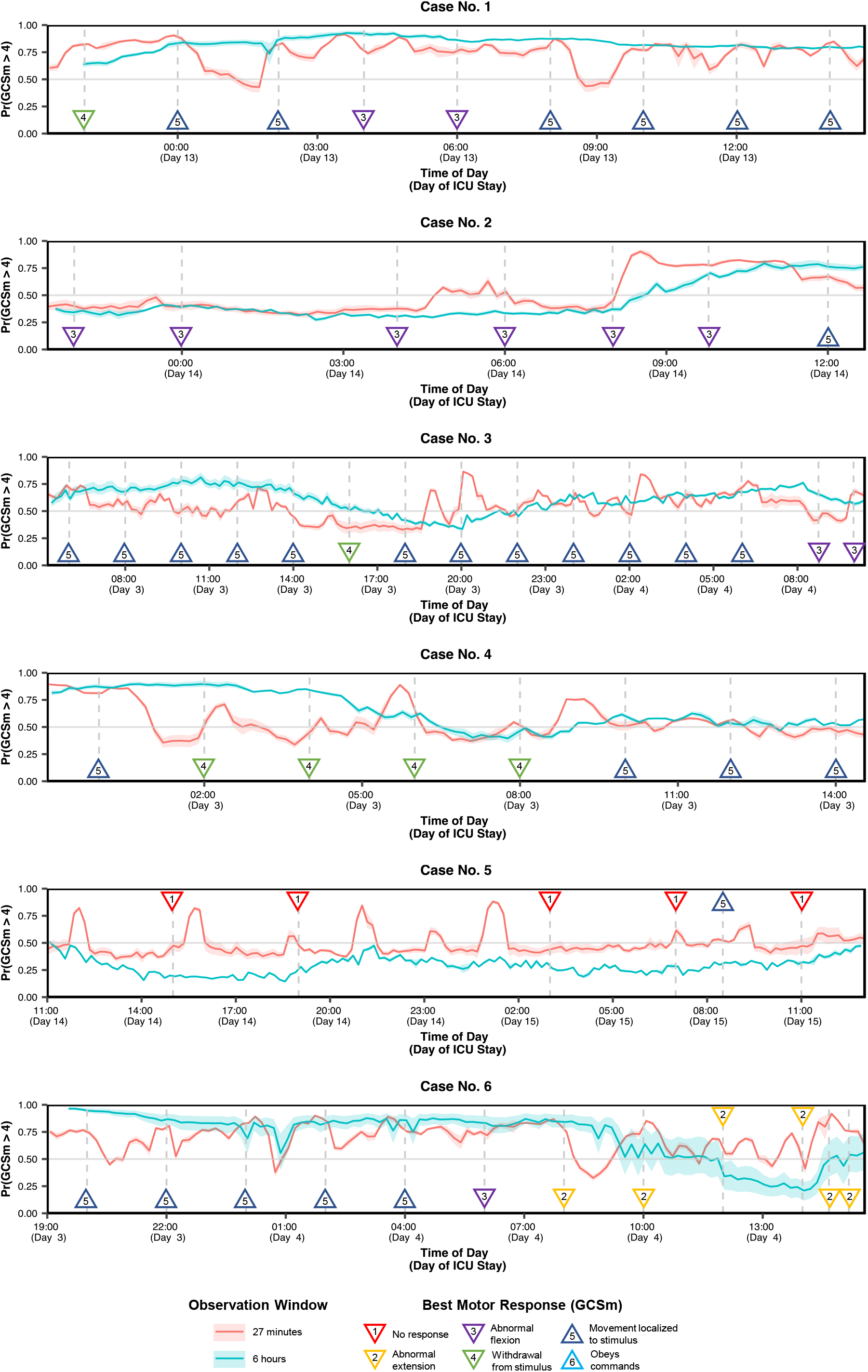
Retrospective case study analysis of accelerometry-based detection of motor function in six patients who experienced relevant transition. The red and blue lines correspond to the predicted probabilities returned every 10 minutes by models trained on all other patients on short (27 minutes) and long (6 hours) observation windows respectively. The predictions from the shorter observation window (red line) respond quicklier to transient changes in the motor component score of the Glasgow Coma Scale (GCSm) (e.g., Case No. 2) while the predictions from the longer observation window (blue line) responds with greater stability to persistent GCSm transitions (e.g., Case No. 6). Shaded areas are 95% confidence intervals derived using bootstrapping (10,000 resamples) to represent the variation across nine missing value imputations. Upward triangle markers designate GCSm > 4 while downward triangle markers designate GCSm ≤ 4.

In case no. 2, we observed that both model types detect an upward transition in GCSm more than three hours before it was recorded clinically. Likewise, the 27-min observation window model detected a downward transition in GCSm about an hour before the upcoming evaluation in case no. 4 and about two hours before in case no. 3. In cases no. 3, 4, and 6, we observed that the 6-hour observation window detects the appropriate transition in GCSm, but with a delay of 3–6 hours. In cases no. 1 and 5, in which we observe a shift and resettlement of GCSm within a 3–5-hour span, the 6-hour model fails to detect the transition while the 27-min model uncertainly oscillates above and below the midline. In general, the shorter observation window model was more dynamic and detected GCSm transitions faster than the longer observation window model. However, persistent transitions, such as the one observed in case no. 6, were detected with greater stability and reliability by the longer observation window model.

## DISCUSSION

### Key findings

We introduce an accelerometry-based based system in critically ill SBI patients that quantitatively captures multisegmental motor patterns correlating with clinical scores of motor responsiveness and functional outcome. The results reveal a significant (AUC = 0.70 [95% CI: 0.53–0.85]), consistent (observation windows: 12 min – 9 hours) association between extracted motion features and the discrimination of SBI patients capable of purposeful movement (GCSm > 4) and those who are not (GCSm ≤ 4) (**Fig. 2a**). A significant discrimination of purposeful movement was achieved with only 12 minutes of accelerometry recording (**Fig. 2b**), and reliable calibration (**Supplementary Fig. S6 online**) and informative classification (**Table 2**) for GCSm > 4 detection suggest that iterations of this system could be clinically useful in automating motor function monitoring. In case studies (**Fig. 5**), we demonstrate that accelerometry-based systems may detect transitions in motor function up to five hours before a clinical evaluation. We also find that the recommended observation window depends on clinical preference: the shorter (27-minute) observation window model reacts more quickly to daily GCSm transitions while the longer (6-hour) observation window model reacts more reliably to persistent transitions and achieves better overall discrimination performance.

The utility of accelerometry-based features for functional outcome prognosis remains unclear. While we found no signal between motion features and long-term (12 months post discharge) outcomes (**Supplementary Fig. S5 online**), the models accurately predicted functional status at hospital discharge (AUC = 0.82 [95% CI: 0.75–0.90]) at a cutoff of GOSE > 5 vs GOSE ≤ 5 for favorable vs unfavorable outcome (**Fig. 3a**). Patients with GOSE > 5 have upper moderate disability or good recovery and are generally able to resume work or previous activities. However, given the small number of SBI patients with GOSE > 5 at hospital discharge, further validation is necessary to determine the reliability of this result. Conflicting results between different calibration metrics (**Supplementary Table S7 online**) underline the class imbalance problem of GOSE > 5 in our dataset; at the same time, we find the consistent discrimination (**Fig. 3b**) and difference in outcome distribution (**Supplementary Fig. S4 online**) as promising markers for further exploration.

Finally, our analysis of feature significance (**Fig. 4**) reveals that both time-domain and frequency-domain features are important for motor function detection and functional outcome prediction. While sensors placed on the wrist achieved the greatest significance scores overall, particularly for features in the frequency-domain, multisegmental motion capture was validated by comparable significance scores of elbow- and ankle-placed sensors across the feature set.

### Relationship with previous studies and future implications

Results presented here represent, to our knowledge, the first approach to relate quantitative motion time series data to neurological states in SBI patients admitted to the ICU. Activity classification with accelerometry-based features has become widespread outside of the clinical domain, especially with advancements in MEMS technology, machine learning, and data sharing [20,33–35]. However, applications to intrahospital care, particularly intensive care, have been limited [27,36] and have largely taken only simple, threshold-based feature approaches to grossly evaluate motor activity (e.g., actigraphy) [37–40]. Reported success in these studies has been variable, but none of them have combined the high-resolution time-domain, feature-domain, and wavelet-domain analysis found in more recent healthy activity classification studies. The focus of our approach, on the relationship between motor profiles of SBI patients over extended periods of time and clinically relevant neurological states, is novel. It builds upon the developments in time-series analysis, dimensionality reduction, and supervised machine learning from activity classification projects as well as the hypotheses of the clinical validity and utility of accelerometry from applied, medical projects.

A continuous high-frequency motion capture system in the intensive care setting produces a high-volume dataset that is also valuable for data-driven research projects. Profiles of motor activity in SBI are poorly understood and decoding specific features of motion in the time-, frequency-, and wavelet-domains can open a window on internal neurological states. Our results demonstrate both the potential and limitations of accelerometry-based monitoring in the ICU. On one hand, the poor overall performance on short-term (discharge) and long-term (12 month) outcome prediction suggest that our derived motion features may not provide enough reliable information to support integration into dynamic prognosis models or decision support systems for WLST. Conversely, when tied to functional motor states, accelerometry-based features may elucidate fundamental mechanisms underlying the strong association between physical activity and clinical outcomes. We aim to collect more data in the NIMS project and focus our research on the development of motion as a quantitative marker of functional recovery for SBI.

More generally, the intensive care setting is a fertile ground for the development of advanced computational methods and applications of artificial intelligence for monitoring and decision support [41]. Patients are typically interfacing with physiological monitoring systems that generate a large volume of data whose complexity may overwhelm human interpretation alone but may be ideal for the training of analytical systems [42]. Since intensive care specialists typically must make time-sensitive decisions for multiple patients at the same time [43], we expect that a near-real-time computational framework assessing motion features alongside other time-series data continuously could provide valuable decision support. For example, a smart alarm system could continuously monitor motor activity in the ICU, and, in conjunction with other clinical factors, dynamically notify clinicians of changing physiology or therapeutic needs before it is too late. We expect that ongoing and subsequent iterations of this work will enable integration of computational physical activity features into the framework of monitoring, smart alarms, and prognostication in the critical care setting.

### Study limitations

We recognize several limitations in this work that need to be addressed. Our statistical analyses and retrospective validation of GCSm detection and GOSE prediction were performed on a limited sample size (*n* = 69 patients) from a single institution and intensive care facility. Further validation will require repeated trials on larger patient populations across multiple centers. There are also improvements to be done to the sensor itself. The planar dimensions of our currently used accelerometer (42 mm × 32 mm) can be reduced further to increase the resolution of localized motion capture. Furthermore, since accelerometry measurements depend on the orientation of the accelerometer with respect to the vertical (gravitational axis), additional modalities of motor output (i.e., gyroscopy and electromyography) could be integrated into the sensor system to inform computational models on the precise arrangement and neural activation of body segments. This would allow us to derive more physiologically relevant features that correspond to validated models of nervous system injury or disease [10]. We also recommend the development of sensors with higher sampling frequencies (≥40 Hz) to capture extremely fine or fast movements of digits or lower extremities. The clinical data collected in our study does not include potentially significant prognostic factors – e.g., features from brainstem assessments, neuroimaging, invasive brain monitoring, electrophysiological studies including evoked potentials, serum biomarkers, as well as therapeutic interventions (sedation, mechanical ventilation, administration of neuromuscular blocking agents or catecholaminergic agents) – that may be impactful in SBI recovery. Inclusion of these variables is likely to improve model performance and precision while also accounting for medical interventions that are likely to have an independent effect on physical activity. Additionally, GCSm itself has been criticized for lack of standardization among practitioners [44,45]. GCSm scores for this work were extracted automatically from EHR and were measured from multiple practitioners across the Johns Hopkins Hospital Neurosciences Critical Care Unit (NCCU) staff. Additionally, we did not record patient turning events, which are part of standardized nursing practices implemented in the ICU treatment of comatose and sedated patients. Moving forward, we aim to supplement clinical validation of the motion features with multifactorial associations with other consciousness, functional, cognitive, psycho-behavioral, symptomatic, and social outcome scales of SBI patients [46].

## METHODS

### Study population and experimental protocol

This work was conducted with approval from the Johns Hopkins Medicine Institutional Review Board (IRB00135674) and written informed consent from patients or surrogates. We prospectively enrolled 72 patients admitted to the NCCU who met the following criteria: age ≥ 18 years, SBI defined as an acute brain injury or illness resulting in impaired consciousness, absence of injuries or lesions involving the extremities, and not expected to die or have WLST in the 24 hours following enrollment. Three of these patients were excluded from the study due to withdrawal of consent (*n* = 2) or corruption of accelerometry data during upload to cloud storage (*n* = 1) (**Supplementary Fig. S1 online**).

Patients were evaluated daily while in the NCCU, at hospital discharge, and at 12 months post discharge by research team members. All GCS evaluations during each patient’s hospital stay were automatically extracted from the institutional EHR system (Epic Systems, Madison, WI, USA). For patients who survived through hospital stay (*n* = 53), outcome scores at discharge were calculated by completing GOSE questionnaires [47] based off the neurological exam information on EHR discharge reports. At 12 months (±1 month) after hospital discharge, we were able to reach 36 patients or carers by telephone and calculated GOSE scores either by performing a validated questionnaire [47] (*n* = 28) or by being notified of the patient’s death (*n* = 8). We confirmed the death (within 12 months of hospital discharge) of 4 additional patients by matching their information with national obituary records. For 8 additional patients, we were able to find EHR notes from a follow-up clinical visit 11 – 13 months after hospital discharge, and we were able to complete a GOSE questionnaire [47] from their neurological exam results. Thus, we lost 5 patients to follow-up (53 – 36 – 4 – 8 = 5), and, including the 16 patients who died during hospital stay, our 12-month post-discharge sample size was *n* = 64. This information is also visualized as a flow diagram in **Supplementary Figure S1 online**.

From the first 3 patients, we collected 10 hours of continuous triaxial accelerometry data, and for the remainder of the patients, we augmented our intended recording duration to between 24 and 48 hours.

### Instrumentation for accelerometry capture

Triaxial sensors (SensorTags CC2650, Texas Instruments, Dallas, TX, USA) were attached with transparent film dressing (Tegaderm Diamond Pattern 1686, 3M, Maplewood, MN, USA) bilaterally near the joints (with standardized orientation) designated in **Fig. 1a**. An additional sensor was placed vertically on the foot of the patient bed to detect patient-independent bed movements. Sensors were equipped with MEMS, variable capacitance tri-axial accelerometers (MPU-9250 MotionTracking Device, TDK InvenSense, San Jose, CA, USA) with sampling frequency (*f*_*s*_) set to 10 Hz, the range of measurable amplitude at ±16 *g* (±157 m/s^2^), and sensitivity at ±4,800 least significant bits per *g* (LSB/*g*).

The sensors transmitted data via a 2.4-GHz Bluetooth antenna to a portable Linux computer (RPi 3 Model B, Raspberry Pi Foundation, Cambridge, UK) placed in the NCCU room. We would execute a Python script on the computer to collect 3 channels (axes) of accelerometry time series from each of the 7 active accelerometers in parallel. The system would log interruptions on a separate .txt file in the instance of a sensor failure. During each trial, we also recorded a video stream (M1045-LW Network Camera, Axis Communications, Lund, Sweden) of the patient that clearly shows the location of each sensor. In the event of sensor interruptions, irregular movement profiles, or bed-sensor-extracted signal magnitude area (SMA) values above 0.135 *g* [32], we would check the footage to identify the source of these results.

### Accelerometry processing and motion feature extraction

Each axial component of each sensor was convolved with a 4^th^-order Butterworth high-pass filter with a critical frequency of *f*_*c*_ = 0.2 Hz (**Supplementary Fig. S10 online**) to remove the baseline offset of accelerometry readings (**Fig. 1a**) and generally separate the low frequency effect of static orientation from the high frequency effect of active body movement [48].

Filtered time-series were segmented into non-overlapping 5-second windows (∼50 data points per window) for motion feature extraction. We selected the motion features listed in **Table 3**, which performed well in physical activity classification tasks [20], to represent three different domains (time, frequency, and wavelet). PDA is defined as the proportion of SMA over 0.135 *g* for each sensor in an observation window (**Fig. 1b**). The remaining features are defined by the following formulae for each 5-second window:

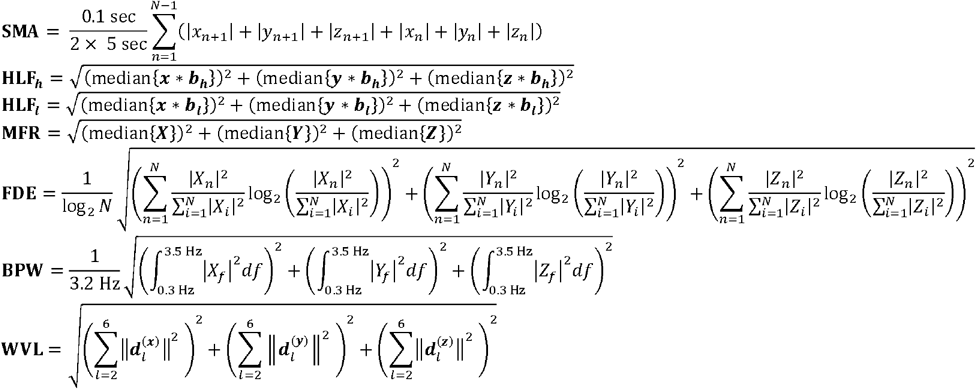

where:

⍰ *x, y, z* represent the x-, y- and z-axes vectors, respectively, of the filtered accelerometry time series within the given 5 second window and *x*_*n*_, *y*_*n*_, *z*_*n*_represent the *n*^th^ elements of these vectors.
⍰ *N* represents the length of each of the *x, y, z* vectors.
⍰ * represents the 1-dimensional convolution operator.
⍰ b_*h*_ represents a 1-dimensional, 4^th^ -order high-pass Butterworth filter with *f*_*c*_ = 2.5 Hz.
⍰ b_*l*_ represents a 1-dimensional, 4^th^ -order low-pass Butterworth filter with *f*_*c*_ = 2.5 Hz.
⍰ *X, Y, Z* represent the discrete Fourier transforms of the x, y, z vectors respectively where *X*_*n*_, *Y*_*n*_, *Z*_*n*_ represent the *n*^th^ elements of these Fourier transform vectors and *X*_*f*_, *Y*_*f*_, *Z*_*f*_ represent the coefficients of the Fourier transforms that correspond to linear frequency *f*.
⍰ 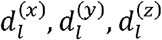 represent the vector of *l*^th^ -level detail coefficients of the 5^th^ –order Daubechies wavelet transform of the *x, y, z* vectors respectively. Post-capture processing of accelerometry were performed offline using MATLAB (Version 9.8.0, MathWorks, Natick, MA, USA) with the Signal Processing, Wavelet, System Identification, and Symbolic Toolboxes.

### Multiple imputation of missing motion features

Due to insufficient battery on the sensors, bedside interventions, interfering equipment, or patient migrations for surgery, imaging, or interunit transfers, a median 1.56% per sensor of each patient’s intended recording duration was missing in our dataset. Missing motion features were multiply imputed (*m* = 9) with a normal (features were normalized with the Box-Cox transform [49]) multivariate time-series algorithm from the ‘Amelia II’ package (v1.7.6) [50] in R (v4.0.0) [51]. The algorithm exploits both spatial correlation (motion feature correlation across the sensors of the same participant) and temporal correlation (autocorrelation structures within each sensor’s time series) to stochastically impute missing time series values in multiple, independently trained runs. We formed subsequent statistical analyses on all 9 imputations to account for variation across imputation.

This model assumes the data is missing at random (MAR) (i.e., the pattern of missingness is independent of unobserved data [52]), which we validated by observing the independence of missingness from sensor placement or time of day (**Supplementary Fig. S11 online**). A complete characterization of the missing data of each patient can be found in **Supplementary Table S8 online**.

### Correction of gross external movements

At time points where the bed-placed sensor SMA exceeded 0.135 *g* (a proposed threshold between static and dynamic activity [32]) and preceded a spike in extremity feature values (1.33% of the time), the bed sensor values of SMA, HLF, BPW, and WVL were subtracted from the extremity values and the bed sensor values of MFR and FDE were added to the extremity values. If a resulting correction value ended up out of a feasible range of static activity for the feature, we replaced the value with a random value, selected uniformly from the static activity range of that feature (**Supplementary Table S9 online**).

### Repeated *k*-fold cross-validation for unbiased model validation

The study population (*n* = 69) was partitioned 25 times with repeated *k*-fold cross-validation (5 repeats, 5 folds) into training sets (∼80%, *n* ≈ 55) and validation sets (∼20%, *n* ≈ 14) for each of the 19 tested observation windows (**Supplementary Table S1 online**) for each of the three tested endpoints (**Fig. 2b**). In splits for motor function detection, patients were stratified by median GCSm over their available observations, while in splits for functional outcome detection, patients were stratified by GOSE scores. One of the nine missing value imputations was drawn with replacement for each partition.

Repeated cross-validation partitions were performed with the ‘caret’ package (v6.0-86) [53] in R.

### Motor function detection

We tested 19 unique observation window durations, from 3 minutes to 24 hours, (**Supplementary Table S1 online**) of accelerometry-derived features directly preceding GCSm evaluations (**Fig. 1b**) at any point during ICU stay. At each of these evaluation points, motion features were organized into matrices where each column represents a unique combination of motion feature type (8 total), sensor placement (6 total), and, for non-PDA features, time before the evaluation. Columns were normalized based on distributions of each placement-feature type combination (48 in total) in the training set. Normalized matrices underwent supervised dimensionality reduction with linear optimal low-rank projection (LOL) [54] learned from the training set. Target dimensionality (*d* ∈ [2,20]) was tested as a model hyperparameter. Low-dimensional vectors of each *d* then underwent element-wise Yeo-Johnson transforms [55] for scaled normalization (learned from the training set) and were used to train and validate logistic regression (‘glm’) models with binary endpoints at each GCSm threshold. All these steps were performed in R.

### Functional outcome prediction

The methodology for functional outcome prediction was identical to that of motor function detection except that GOSE thresholds instead of GCSm thresholds were used as endpoints.

### Assessment of model performance and calibration on validation sets

Both motor function detection and functional outcome prediction models were trained and validated on each of the 25 repeated cross-validation splits for each of the 19 observation windows for each of the 19 unique target dimensionalities (*d*) for each of the endpoint thresholds (5 for GCSm, 5 for GOSE at discharge, 7 for GOSE at 12 months). Models returned binary prediction probabilities as well as a classification based on a probability threshold of 0.5 for each validation set observation.

Based on the validation set predictions, we calculated metrics of binary outcome discrimination performance (**Supplementary Tables S2, S4**, and **S6 online**), classification performance (**Table 2**), and probability calibration [31,56] (**Table 3**). We also visualized ROC curves (**Fig. 2a, 3a**, and **Supplementary Fig. S5 online**), probability calibration curves (**Supplementary Fig. S6 online**), and, in one case, the precision recall curve (**Supplementary Fig. S4 online**) of the optimally discriminating (maximal AUC) models to assess discrimination, calibration, and case detection power respectively. We calculated unbiased mean values and 95% confidence intervals for both metrics and curves with bootstrap bias-corrected cross-validation (BBC-CV) with repeats [57] on 1,000 resamples of the patient set across the validation set predictions. In this way, 95% confidence intervals account for the variation across the patient set, across the nine missing value imputations, and across the 25 repeated cross-validation partitions.

### Feature significance scores

The coefficients (i.e., loadings) of the trained LOL projection matrix represent the relative importance of each column in explaining the variance in the dataset stratified by the endpoint [54]. Thus, we derived a relative importance score of each sensor-feature type combination for both motor function detection and functional outcome prediction by multiplying the mean absolute value of the loadings per each combination and the absolute value of the trained logistic regression coefficient of the corresponding reduced dimension. This would be performed across all 25 partitions of each combination of observation window, threshold, and endpoint. We then calculated 95% confidence intervals on feature significance scores by bootstrapping 1,000 resamples across the 25 repeated cross-validation folds and nine missing value imputations.

### Ethics and informed consent statement

Informed consent was obtained from all participants or legally identified surrogates in this study and the procedure was approved by the Institutional Review Board of the Johns Hopkins Medicine Institutional Review Board (reference number: IRB00135674). All research was performed in accordance with relevant guidelines and regulations.

## Data Availability

Per our current Johns Hopkins Medicine IRB protocol (IRB00135674), we are not permitted to share the clinical data collected for this study. However, we welcome all forms of collaboration, and urge interested investigators to contact the corresponding author (SB) with their institutional affiliation and proposed use of the dataset to submit a new protocol for access. The data may not be used for commercial products or redistributed in any way.

## CODE AVAILABILITY

All code used in the data collection and analyses outlined in this manuscript can be found at the following GitHub repository [58]: https://github.com/sbhattacharyay/nims (DOI: 10.5281/zenodo.4765305).

## ACKNOWLEDGEMENTS

We graciously acknowledge the patients, families, NCCU nurses, and physicians who participated in and contributed to this study. S.B. would like to thank Kathleen Mitchell-Fox (Univ. of Cambridge) for reviewing and offering comments on the manuscript. We also wish to specifically thank Aditya Joshi (Rowan Univ.), Sanya Yadav (Univ. of Pittsburgh), Tobias Fauser (Univ. of Arizona), Michiru Fredricks (Johns Hopkins Univ.), Alexander Sigmon (Johns Hopkins Univ.), Shikha Gandhi (Johns Hopkins Univ.), and Joshua Vogelstein (Johns Hopkins Univ.) for their roles in the early development, data curation, and advising of statistical methodologies of the NIMS project.

This work was partially supported by awards from the Johns Hopkins University Office of the Provost and the Hodson Trust, received by S.B. S.B. is currently funded by a Gates Cambridge fellowship.

## AUTHOR CONTRIBUTIONS

S.B. co-conceptualized the study, developed the methodology of the experiments, acquired accelerometry data from patients, acquired funding for the project, performed statistical analyses on the data, visualized the results for publication, and wrote the complete manuscript. J.R. and R.E.C. aided in the conceptualization and data collection of this work and revised the manuscript. M.W., H.B.K., and E.J. aided S.B. in the statistical analysis, processing of data, and visualization of results. P.H.D. extracted neurological assessment scores from electronic health records. E.C. recruited patients for the study, performed clinical surveys, and collected clinical data from patient records. P.K. aided in the conceptualization of the study and the development of the methodology and established the data acquisition infrastructure.

R.D.S. served as the principal investigator, co-conceptualized the study, aided in the development of the methodology, procured IRB approval for data collection from human subjects, aided in data collection, provided access to clinical resources at the Johns Hopkins Hospital, and revised the manuscript.

