## Supplementary Information for "Decoding accelerometry for classification and prediction of critically ill patients with severe brain injury"

**The file includes:**


| | Description | Page(s) |
|---|---|---|
| **Fig. S1** | CONSORT-style flow diagram for patient enrollment and follow-up | 2 |
| **Fig. S2** | Count histograms of accelerometry recording information | 3 |
| **Fig. S3** | Trajectories of motor component scores of the Glasgow Coma Scale (GCSm) of each study participant during ICU stay | 4–7 |
| **Table S1** | Count distributions of GCSm scores per observation window | 8 |
| **Table S2** | Discrimination of threshold-level GCSm detection models per observation window | 9 |
| **Table S3** | Count distributions of GOSE scores at hospital discharge per observation window | 10 |
| **Table S4** | Discrimination of threshold-level GOSE at hospital discharge prediction models per observation window | 11 |
| **Fig. S4** | Precision recall curve and prediction distribution of GOSE (discharge) > 5 prediction | 12 |
| **Table S5** | Count distributions of GOSE scores at 12 months post discharge per observation window | 13 |
| **Fig. S5** | Discrimination performance of functional outcome at 12 months post discharge prediction models on validation sets | 14 |
| **Table S6** | Discrimination of threshold-level GOSE at 12 months post discharge prediction models per observation window | 15 |
| **Fig. S6** | Probability calibration of optimally discriminating motor function detection and functional outcome prediction models on validation sets | 16–17 |
| **Table S7** | Probability calibration metrics of optimally discriminating models | 18 |
| **Fig. S7** | Mean motion feature trajectories in the six hours preceding GCSm evaluation, stratified by GCSm scores and bilateral sensor placement | 19 |
| **Fig. S8** | Correlation matrices of extracted motion features across different sensor placements | 20 |
| **Fig. S9** | Violin plots of extracted motion feature values (30 min observation window), stratified by bilateral sensor placement and GCSm scores | 21 |
| **Fig. S10** | Bode plot of filter used in accelerometry processing | 22 |
| **Fig. S11** | Percentages of missing, static, and dynamic accelerometry data by time of day of recording and sensor placement | 23 |
| **Table S8** | Percentages of missing accelerometry data per sensor and recording duration of each study participant | 24 |
| **Table S9** | Ranges of static activity values for each motion feature | 25 |

### Supplementary Figure S1

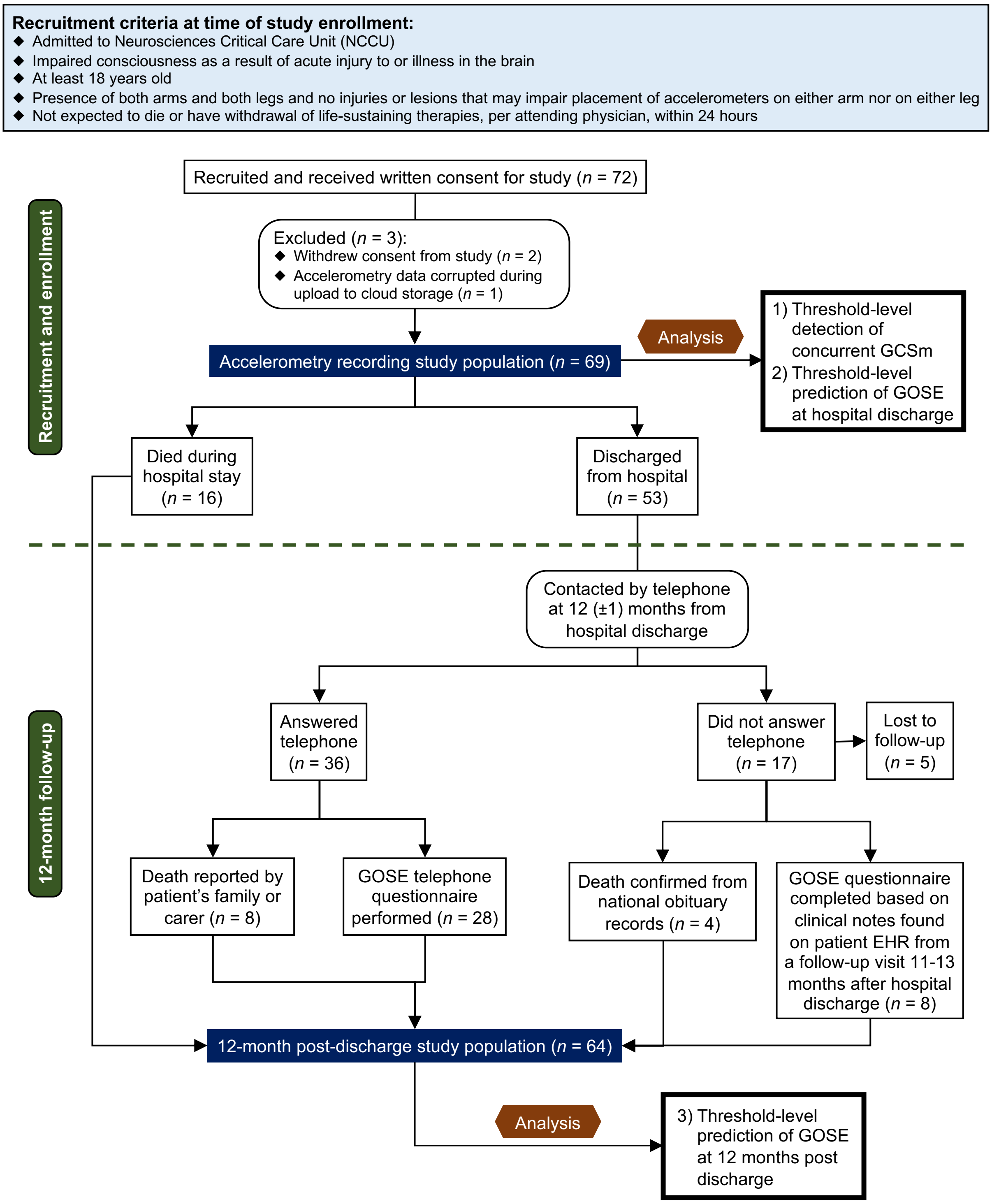


**Fig. S1. CONSORT-style flow diagram for patient enrollment and follow-up.** The dashed, olive-green line in the upper-middle of the diagram represents the time of discharge from the hospital. Abbreviations: motor component score of the Glasgow Coma Scale (GCSm), Glasgow Outcome Scale – Extended (GOSE).

**Supplementary Figure S2**

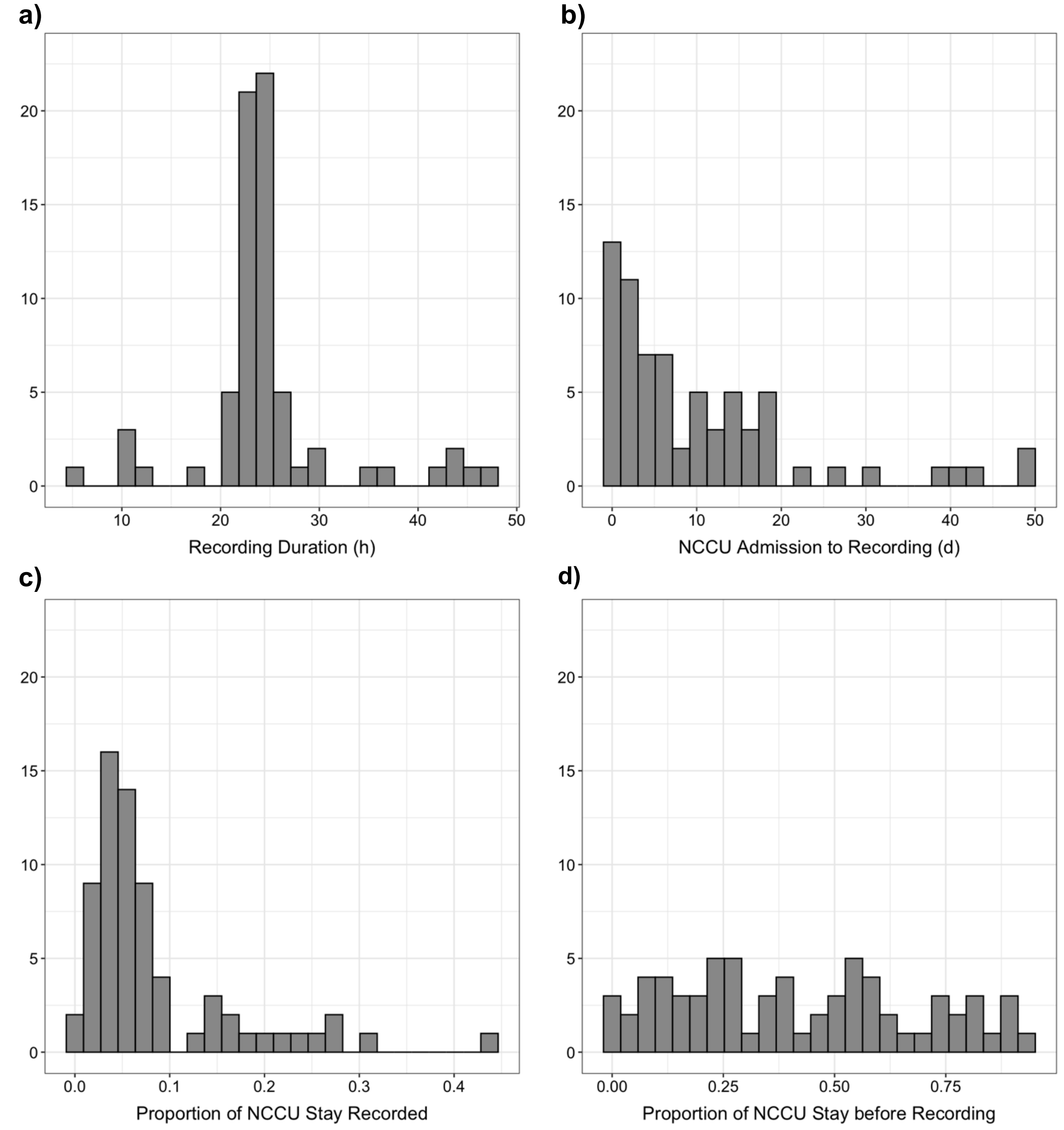


**Fig. S2. Count histograms of accelerometry recording information.** The histograms (*n* = 69, 25 uniform partitions) display the distributions of (**a**) accelerometry recording duration in hours, (**b**) delay between ICU admission and start of accelerometry recording in days, (**c**) proportion of ICU stay during which accelerometry recording took place, and (**d**) proportion of ICU stay that elapsed before accelerometry recording began.

### **Supplementary Figure S3** (*part 1 of 3*)

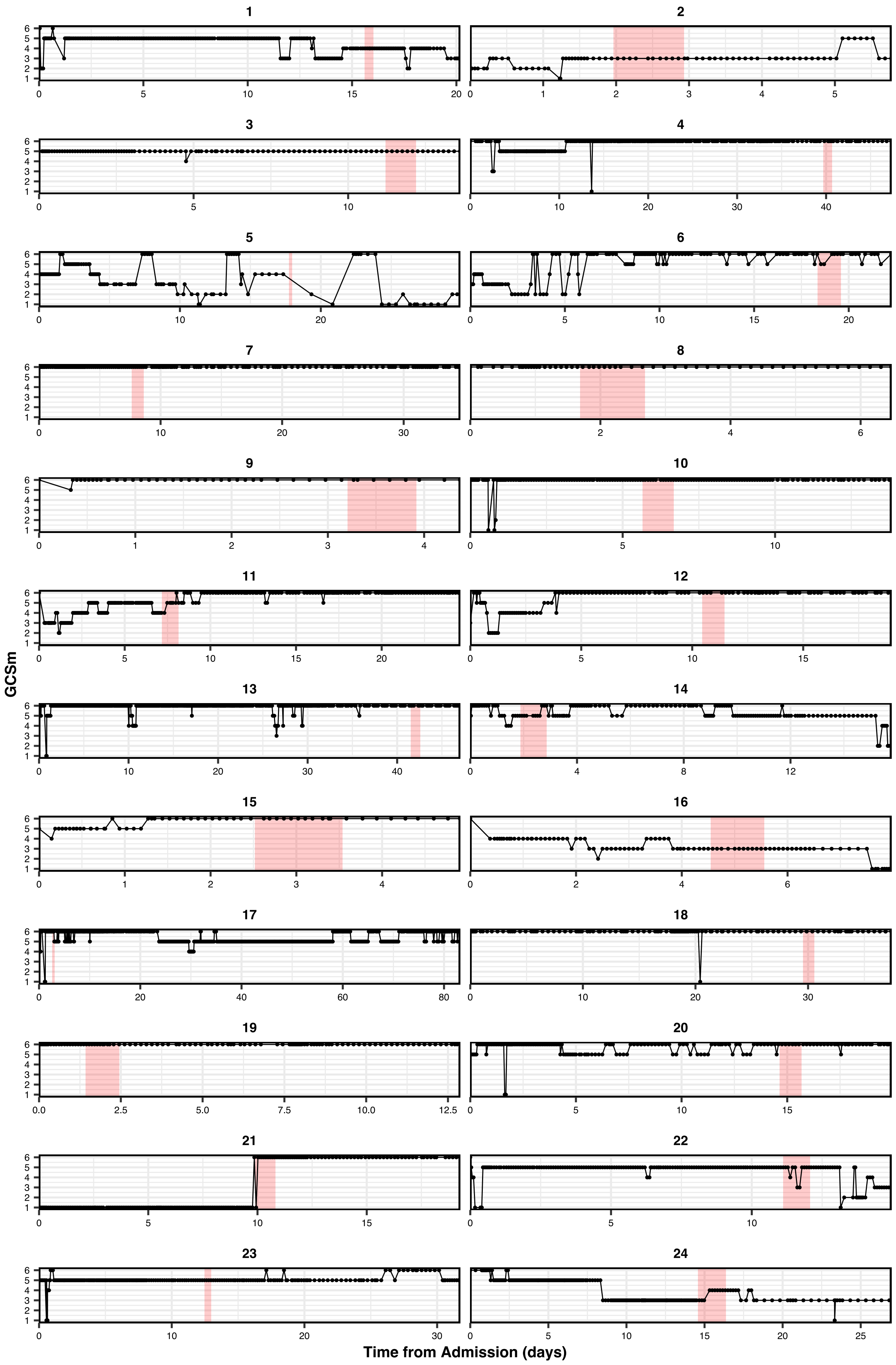

Supplementary Figure S3 continued (*part 2 of 3*)

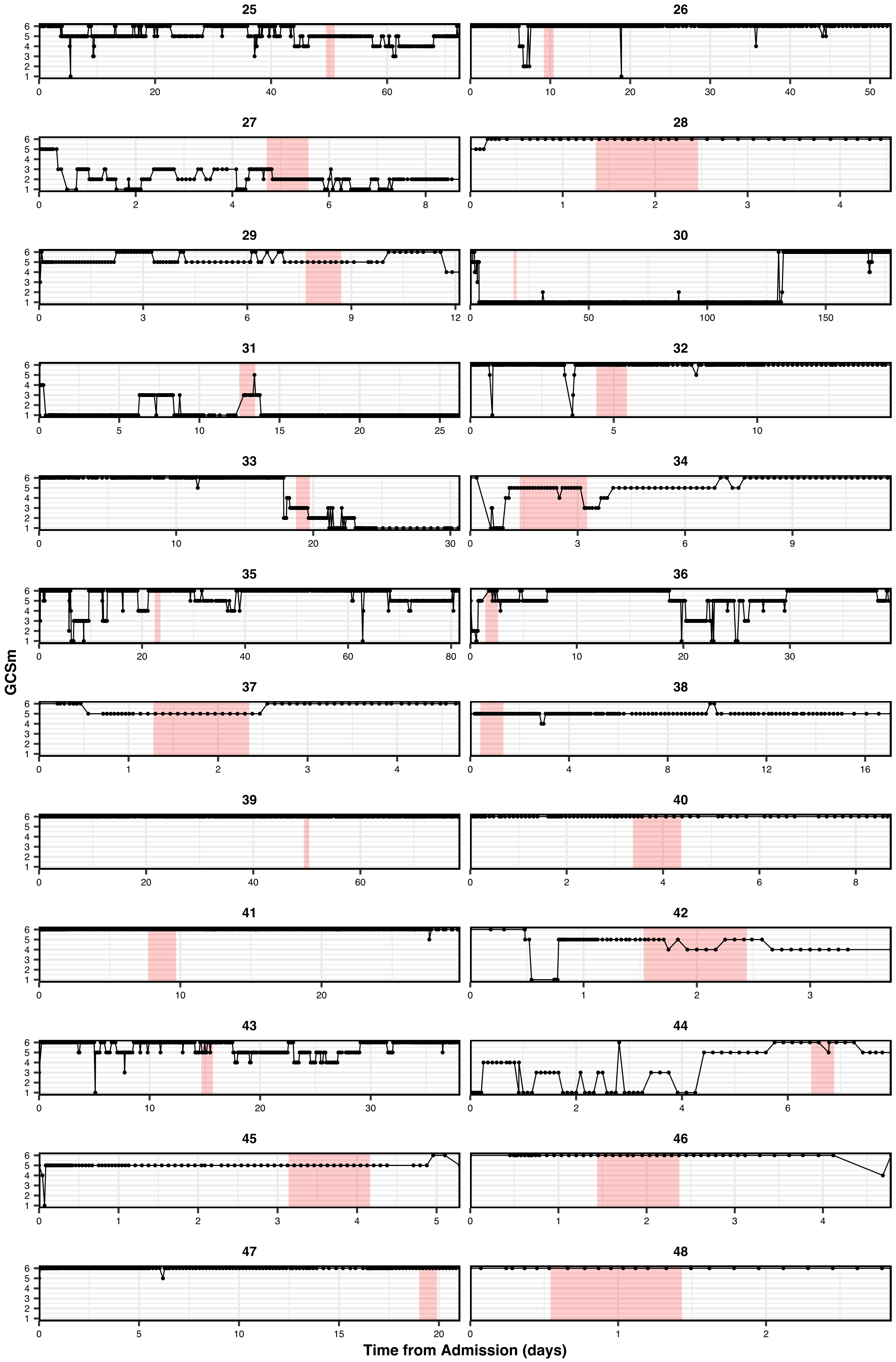

Supplementary Figure S3 continued (*part 3 of 3*)

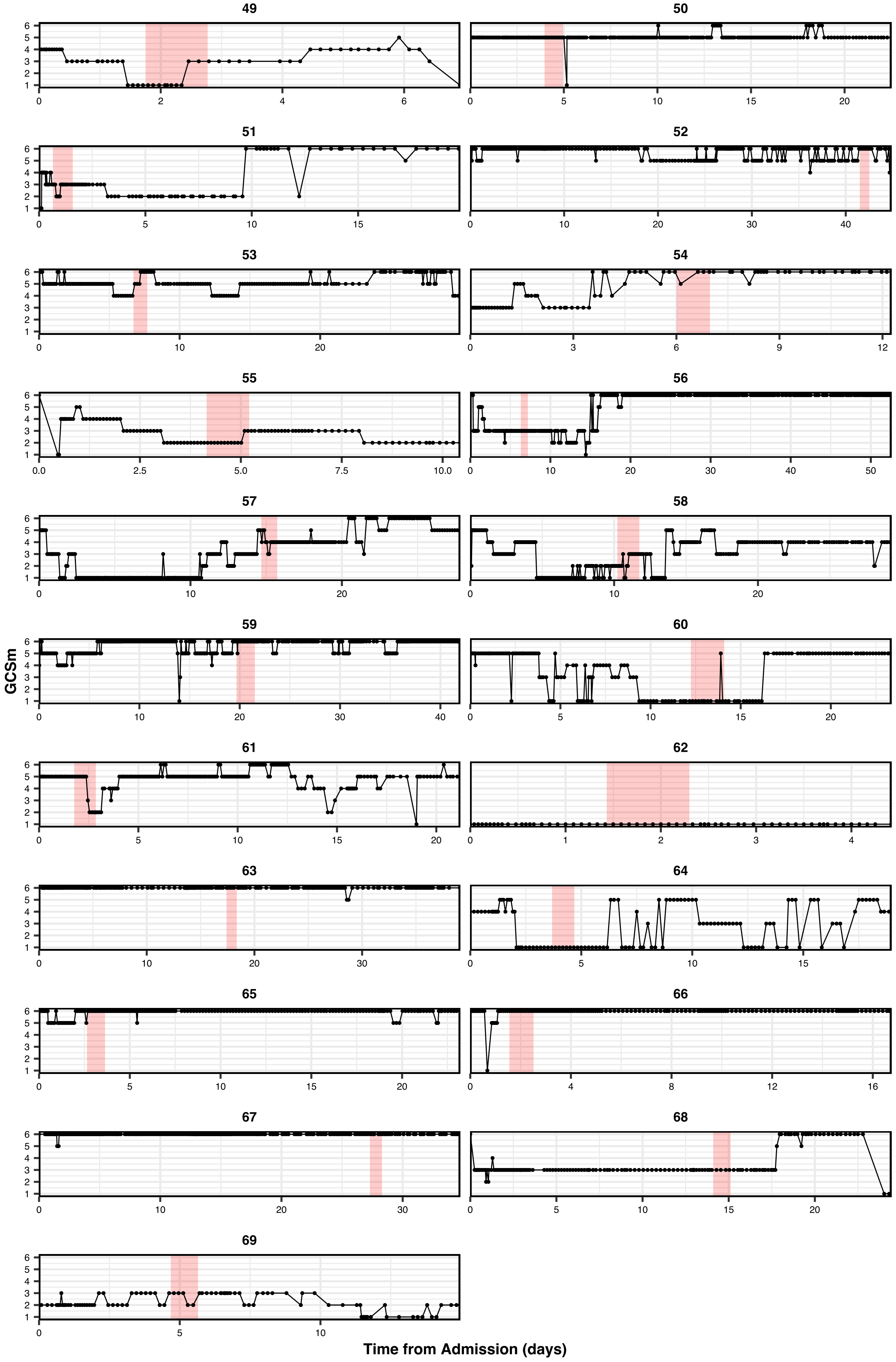

**Fig. S3. Trajectories of motor component scores of the Glasgow Coma Scale (GCSm) of each study participant during ICU stay.** Shaded areas represent time ranges during which we recorded accelerometry from the corresponding patient and points mark the exact times of a GCSm evaluation.

**Supplementary Table S1**: Count distributions of GCSm scores per observation window.

| Observation Window (hr) | Unique patients (*n*) | Total observations (*n*) | Motor component score of the Glasgow Coma Scale (GCSm) | | | | | |
|---|---|---|---|---|---|---|---|---|
| | | | 1 | 2 | 3 | 4 | 5 | 6 |
| 0.05 | 68 | 649 | 42 (6.47%) | 51 (7.86%) | 98 (15.10%) | 30 (4.62%) | 154 (23.73%) | 274 (42.22%) |
| 0.1 | 68 | 648 | 42 (6.48%) | 51 (7.87%) | 98 (15.12%) | 30 (4.63%) | 154 (23.77%) | 273 (42.13%) |
| 0.15 | 68 | 647 | 42 (6.49%) | 51 (7.88%) | 98 (15.15%) | 30 (4.64%) | 154 (23.80%) | 272 (42.04%) |
| 0.2 | 68 | 645 | 42 (6.51%) | 51 (7.91%) | 98 (15.19%) | 30 (4.65%) | 154 (23.88%) | 270 (41.86%) |
| 0.25 | 68 | 644 | 42 (6.52%) | 51 (7.92%) | 98 (15.22%) | 30 (4.66%) | 153 (23.76%) | 270 (41.93%) |
| 0.3 | 68 | 642 | 42 (6.54%) | 51 (7.94%) | 98 (15.26%) | 30 (4.67%) | 153 (23.83%) | 268 (41.74%) |
| 0.35 | 68 | 642 | 42 (6.54%) | 51 (7.94%) | 98 (15.26%) | 30 (4.67%) | 153 (23.83%) | 268 (41.74%) |
| 0.4 | 68 | 641 | 42 (6.55%) | 51 (7.96%) | 98 (15.29%) | 30 (4.68%) | 153 (23.87%) | 267 (41.65%) |
| 0.45 | 68 | 640 | 42 (6.56%) | 51 (7.97%) | 98 (15.31%) | 30 (4.69%) | 152 (23.75%) | 267 (41.72%) |
| 0.5 | 68 | 636 | 42 (6.60%) | 50 (7.86%) | 97 (15.25%) | 29 (4.56%) | 151 (23.74%) | 267 (41.98%) |
| 1 | 68 | 624 | 41 (6.57%) | 50 (8.01%) | 93 (14.90%) | 29 (4.65%) | 148 (23.72%) | 263 (42.15%) |
| 2 | 68 | 599 | 39 (6.51%) | 49 (8.18%) | 87 (14.52%) | 28 (4.67%) | 141 (23.54%) | 255 (42.57%) |
| 3 | 68 | 564 | 37 (6.56%) | 47 (8.33%) | 82 (14.54%) | 28 (4.96%) | 130 (23.05%) | 240 (42.55%) |
| 6 | 68 | 488 | 32 (6.56%) | 38 (7.79%) | 73 (14.96%) | 23 (4.71%) | 109 (22.34%) | 213 (43.65%) |
| 9 | 66 | 405 | 26 (6.42%) | 30 (7.41%) | 64 (15.80%) | 19 (4.69%) | 90 (22.22%) | 176 (43.46%) |
| 12 | 64 | 334 | 20 (5.99%) | 26 (7.78%) | 49 (14.67%) | 16 (4.79%) | 72 (21.56%) | 151 (45.21%) |
| 15 | 63 | 259 | 15 (5.79%) | 20 (7.72%) | 37 (14.29%) | 14 (5.41%) | 57 (22.01%) | 116 (44.79%) |
| 18 | 62 | 190 | 11 (5.79%) | 14 (7.37%) | 26 (13.68%) | 11 (5.79%) | 40 (21.05%) | 88 (46.32%) |
| 24 | 16 | 62 | 4 (6.45%) | 3 (4.84%) | 9 (14.52%) | 7 (11.29%) | 14 (22.58%) | 25 (40.32%) |

**Supplementary Table S2**: Discrimination of threshold-level GCSm detection models per observation window.

| Observation Window (hr) | Unique patients (*n*) | Total observations (*n*) | AUC at different motor component score of the Glasgow Coma Scale (GCSm) thresholds | | | | |
|---|---|---|---|---|---|---|---|
| | | | GCSm > 1 | GCSm > 2 | GCSm > 3 | GCSm > 4 | GCSm > 5 |
| 0.05 | 68 | 649 | 0.54 (0.09–0.73) | 0.49 (0.37–0.61) | 0.55 (0.46–0.65) | 0.59 (0.50–0.69) | 0.56 (0.47–0.64) |
| 0.1 | 68 | 648 | 0.52 (0.26–0.71) | 0.51 (0.34–0.66) | 0.56 (0.47–0.66) | 0.60 (0.50–0.70) | 0.59 (0.50–0.67) |
| 0.15 | 68 | 647 | 0.49 (0.31–0.64) | 0.48 (0.33–0.61) | 0.56 (0.46–0.67) | 0.61 (0.50–0.72) | 0.56 (0.47–0.66) |
| 0.2 | 68 | 645 | 0.54 (0.31–0.73) | 0.50 (0.36–0.63) | 0.58 (0.48–0.69) | 0.65 (0.54–0.75) | 0.55 (0.46–0.63) |
| 0.25 | 68 | 644 | 0.53 (0.31–0.67) | 0.56 (0.39–0.68) | 0.56 (0.46–0.65) | 0.63 (0.51–0.76) | 0.56 (0.47–0.65) |
| 0.3 | 68 | 642 | 0.56 (0.25–0.82) | 0.52 (0.38–0.65) | 0.61 (0.50–0.73) | 0.66 (0.54–0.78) | 0.58 (0.48–0.67) |
| 0.35 | 68 | 642 | 0.57 (0.26–0.80) | 0.57 (0.38–0.75) | 0.60 (0.48–0.72) | 0.66 (0.53–0.79) | 0.55 (0.46–0.63) |
| 0.4 | 68 | 641 | 0.53 (0.35–0.71) | 0.55 (0.38–0.67) | 0.60 (0.47–0.72) | 0.67 (0.53–0.79) | 0.58 (0.48–0.66) |
| 0.45 | 68 | 640 | 0.54 (0.30–0.66) | 0.52 (0.38–0.65) | 0.63 (0.50–0.76) | 0.68 (0.55–0.80) | 0.59 (0.51–0.68) |
| 0.5 | 68 | 636 | 0.58 (0.16–0.79) | 0.53 (0.35–0.69) | 0.60 (0.48–0.73) | 0.62 (0.51–0.72) | 0.58 (0.48–0.66) |
| 1 | 68 | 624 | 0.56 (0.19–0.75) | 0.54 (0.38–0.68) | 0.59 (0.45–0.73) | 0.67 (0.52–0.82) | 0.60 (0.49–0.70) |
| 2 | 68 | 599 | 0.60 (0.13–0.81) | 0.59 (0.38–0.79) | 0.65 (0.51–0.80) | 0.69 (0.53–0.83) | 0.66 (0.53–0.78) |
| 3 | 68 | 564 | 0.67 (0.15–0.87) | 0.60 (0.43–0.80) | 0.61 (0.48–0.74) | 0.69 (0.54–0.85) | 0.63 (0.51–0.74) |
| 6 | 68 | 488 | 0.69 (0.15–0.88) | 0.56 (0.39–0.76) | 0.65 (0.49–0.81) | 0.70 (0.53–0.85) | 0.59 (0.46–0.72) |
| 9 | 66 | 405 | 0.47 (0.34–0.56) | 0.53 (0.38–0.74) | 0.64 (0.46–0.82) | 0.70 (0.50–0.87) | 0.61 (0.48–0.72) |
| 12 | 64 | 334 | 0.41 (0.17–0.51) | 0.48 (0.36–0.60) | 0.59 (0.44–0.75) | 0.63 (0.48–0.80) | 0.58 (0.45–0.71) |
| 15 | 63 | 259 | 0.68 (0.53–0.84) | 0.52 (0.39–0.67) | 0.55 (0.39–0.68) | 0.64 (0.45–0.83) | 0.59 (0.44–0.72) |
| 18 | 62 | 190 | 0.65 (0.53–0.77) | 0.56 (0.39–0.73) | 0.48 (0.36–0.61) | 0.69 (0.48–0.90) | 0.52 (0.37–0.66) |
| 24 | 16 | 62 | | 0.26 (0.10–0.43) | 0.36 (0.12–0.62) | 0.44 (0.11–0.73) | 0.38 (0.10–0.71) |

Values in the five rightmost columns represent mean validation set area under the receiver operating characteristic curve (AUC) values with associated 95% confidence intervals in parentheses. Confidence intervals were derived using bias-corrected bootstrapping (1,000 resamples) and represent the variation across repeated cross-validation folds (5 repeats of 5 folds) and nine missing value imputations. Missing values designate insufficient diversity in endpoint labels to evaluate models of that observation window and threshold combination.

**Supplementary Table S3**: Count distributions of GOSE scores at hospital discharge per observation window.

| Obs. Window (hr) | Unique patients (*n*) | Total obs. (*n*) | Glasgow Outcome Scale – Extended (GOSE) at hospital discharge | | | | | | | |
|---|---|---|---|---|---|---|---|---|---|---|
| | | | 1 | 2 | 3 | 4 | 5 | 6 | 7 | 8 |
| 0.05 | 68 | 649 | 169 (26.04%) | 17 (2.62%) | 265 (40.83%) | 119 (18.34%) | 66 (10.17%) | 10 (1.54%) | 3 (0.46%) | 0 |
| 0.1 | 68 | 648 | 168 (25.93%) | 17 (2.62%) | 265 (40.90%) | 119 (18.36%) | 66 (10.19%) | 10 (1.54%) | 3 (0.46%) | 0 |
| 0.15 | 68 | 647 | 168 (25.97%) | 17 (2.63%) | 264 (40.80%) | 119 (18.39%) | 66 (10.20%) | 10 (1.55%) | 3 (0.46%) | 0 |
| 0.2 | 68 | 645 | 168 (26.05%) | 17 (2.64%) | 264 (40.93%) | 119 (18.45%) | 64 (9.92%) | 10 (1.55%) | 3 (0.47%) | 0 |
| 0.25 | 68 | 644 | 167 (25.93%) | 17 (2.64%) | 264 (40.99%) | 119 (18.48%) | 64 (9.94%) | 10 (1.55%) | 3 (0.47%) | 0 |
| 0.3 | 68 | 642 | 167 (26.01%) | 17 (2.65%) | 263 (40.97%) | 119 (18.54%) | 63 (9.81%) | 10 (1.56%) | 3 (0.47%) | 0 |
| 0.35 | 68 | 642 | 167 (26.01%) | 17 (2.65%) | 263 (40.97%) | 119 (18.54%) | 63 (9.81%) | 10 (1.56%) | 3 (0.47%) | 0 |
| 0.4 | 68 | 641 | 167 (26.05%) | 17 (2.65%) | 263 (41.03%) | 119 (18.56%) | 62 (9.67%) | 10 (1.56%) | 3 (0.47%) | 0 |
| 0.45 | 68 | 640 | 166 (25.94%) | 17 (2.66%) | 263 (41.09%) | 119 (18.59%) | 62 (9.69%) | 10 (1.56%) | 3 (0.47%) | 0 |
| 0.5 | 68 | 636 | 164 (25.79%) | 17 (2.67%) | 261 (41.04%) | 119 (18.71%) | 62 (9.75%) | 10 (1.57%) | 3 (0.47%) | 0 |
| 1 | 68 | 624 | 162 (25.96%) | 17 (2.72%) | 254 (40.71%) | 117 (18.75%) | 61 (9.78%) | 10 (1.60%) | 3 (0.48%) | 0 |
| 2 | 68 | 599 | 152 (25.38%) | 16 (2.67%) | 248 (41.40%) | 111 (18.53%) | 60 (10.02%) | 9 (1.50%) | 3 (0.50%) | 0 |
| 3 | 68 | 564 | 145 (25.71%) | 15 (2.66%) | 233 (41.31%) | 103 (18.26%) | 56 (9.93%) | 9 (1.60%) | 3 (0.53%) | 0 |
| 6 | 68 | 488 | 120 (24.59%) | 13 (2.66%) | 205 (42.01%) | 91 (18.65%) | 50 (10.25%) | 7 (1.43%) | 2 (0.41%) | 0 |
| 9 | 66 | 405 | 101 (24.94%) | 11 (2.72%) | 170 (41.98%) | 79 (19.51%) | 38 (9.38%) | 6 (1.48%) | 0 | 0 |
| 12 | 64 | 334 | 80 (23.95%) | 9 (2.69%) | 144 (43.11%) | 64 (19.16%) | 33 (9.88%) | 4 (1.20%) | 0 | 0 |
| 15 | 63 | 259 | 57 (22.01%) | 8 (3.09%) | 118 (45.56%) | 50 (19.31%) | 23 (8.88%) | 3 (1.16%) | 0 | 0 |
| 18 | 62 | 190 | 34 (17.89%) | 7 (3.68%) | 94 (49.47%) | 36 (18.95%) | 17 (8.95%) | 2 (1.05%) | 0 | 0 |
| 24 | 16 | 62 | 1 (1.61%) | 5 (8.06%) | 41 (66.13%) | 13 (20.97%) | 2 (3.23%) | 0 | 0 | 0 |

**Supplementary Table S4**: Discrimination of threshold-level GOSE at hospital discharge prediction models per observation window.

| Observation Window (hr) | Unique patients (*n*) | Total observations (*n*) | AUC at different Glasgow Outcome Scale – Extended (GOSE) thresholds at hospital discharge | | | | |
|---|---|---|---|---|---|---|---|
| | | | GOSE > 1 | GOSE > 2 | GOSE > 3 | GOSE > 4 | GOSE > 5 |
| 0.05 | 68 | 649 | 0.53 (0.42–0.64) | 0.51 (0.40–0.61) | 0.54 (0.42–0.66) | 0.58 (0.45–0.75) | 0.40 (0.27–0.49) |
| 0.1 | 68 | 648 | 0.48 (0.39–0.57) | 0.50 (0.40–0.59) | 0.53 (0.44–0.62) | 0.54 (0.40–0.72) | 0.30 (0.09–0.49) |
| 0.15 | 68 | 647 | 0.52 (0.43–0.61) | 0.55 (0.45–0.64) | 0.50 (0.41–0.60) | 0.56 (0.43–0.69) | 0.34 (0.15–0.49) |
| 0.2 | 68 | 645 | 0.50 (0.40–0.60) | 0.53 (0.43–0.63) | 0.49 (0.39–0.59) | 0.54 (0.40–0.70) | 0.42 (0.22–0.61) |
| 0.25 | 68 | 644 | 0.52 (0.41–0.63) | 0.54 (0.42–0.66) | 0.50 (0.40–0.60) | 0.60 (0.47–0.75) | 0.36 (0.21–0.49) |
| 0.3 | 68 | 642 | 0.53 (0.43–0.63) | 0.54 (0.44–0.63) | 0.52 (0.42–0.62) | 0.54 (0.42–0.68) | 0.51 (0.47–0.58) |
| 0.35 | 68 | 642 | 0.53 (0.42–0.63) | 0.53 (0.42–0.62) | 0.51 (0.42–0.59) | 0.52 (0.36–0.72) | 0.49 (0.45–0.54) |
| 0.4 | 68 | 641 | 0.53 (0.42–0.65) | 0.55 (0.43–0.66) | 0.51 (0.41–0.59) | 0.56 (0.43–0.65) | 0.50 (0.45–0.55) |
| 0.45 | 68 | 640 | 0.54 (0.43–0.64) | 0.57 (0.45–0.67) | 0.46 (0.38–0.53) | 0.52 (0.41–0.74) | 0.50 (0.45–0.54) |
| 0.5 | 68 | 636 | 0.52 (0.41–0.63) | 0.55 (0.43–0.66) | 0.49 (0.39–0.57) | 0.52 (0.42–0.66) | 0.52 (0.46–0.59) |
| 1 | 68 | 624 | 0.54 (0.42–0.65) | 0.58 (0.44–0.69) | 0.48 (0.39–0.56) | 0.54 (0.40–0.65) | 0.54 (0.45–0.65) |
| 2 | 68 | 599 | 0.52 (0.41–0.61) | 0.54 (0.43–0.64) | 0.48 (0.37–0.55) | 0.49 (0.36–0.66) | 0.77 (0.71–0.84) |
| 3 | 68 | 564 | 0.54 (0.40–0.67) | 0.58 (0.46–0.70) | 0.52 (0.39–0.64) | 0.57 (0.37–0.74) | 0.80 (0.66–0.96) |
| 6 | 68 | 488 | 0.56 (0.42–0.68) | 0.60 (0.46–0.74) | 0.53 (0.40–0.66) | 0.56 (0.38–0.84) | 0.82 (0.75–0.90) |
| 9 | 66 | 405 | 0.56 (0.39–0.71) | 0.59 (0.43–0.75) | 0.51 (0.38–0.65) | 0.44 (0.20–0.63) | |
| 12 | 64 | 334 | 0.51 (0.33–0.67) | 0.61 (0.44–0.77) | 0.52 (0.38–0.67) | 0.52 (0.38–0.77) | |
| 15 | 63 | 259 | 0.47 (0.32–0.58) | 0.53 (0.37–0.68) | 0.53 (0.38–0.68) | 0.45 (0.28–0.64) | |
| 18 | 62 | 190 | 0.50 (0.38–0.61) | 0.55 (0.38–0.69) | 0.49 (0.39–0.60) | 0.51 (0.33–0.70) | |
| 24 | 16 | 62 | | 0.26 (0.08–0.50) | 0.48 (0.18–0.84) | 0.13 (0.03–0.27) | |

Values in the five rightmost columns represent mean validation set area under the receiver operating characteristic curve (AUC) values with associated 95% confidence intervals in parentheses. Confidence intervals were derived using bias-corrected bootstrapping (1,000 resamples) and represent the variation across repeated cross-validation folds (5 repeats of 5 folds) and nine missing value imputations. Missing values designate insufficient diversity in endpoint labels to evaluate models of that observation window and threshold combination.

**Supplementary Figure S4**

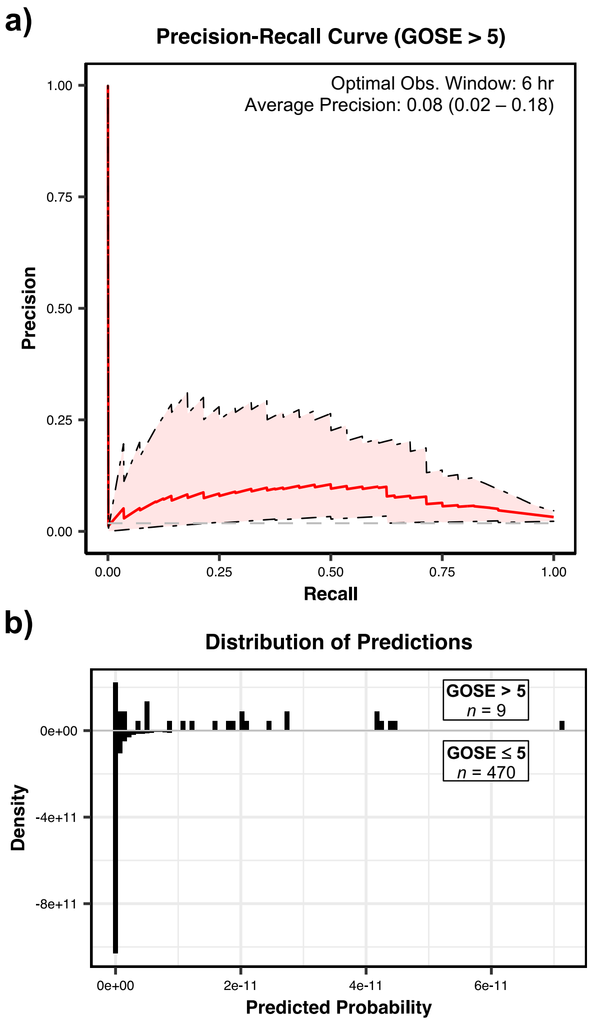


**Fig. S4. Precision recall curve and prediction distribution of GOSE (discharge) > 5 prediction. a** Precision recall curve of optimally discriminating model configuration of GOSE > 5 prediction at hospital discharge (**Fig. 3a**). Shaded areas represent 95% confidence intervals derived using bias-corrected bootstrapping (1,000 resamples) to represent the variation across repeated cross-validation folds (5 repeats of 5 folds) and nine missing value imputations. The values in the box represent the observation window of the optimally discriminating model as well as the mean average precision (with 95% confidence interval in parentheses). The horizontal dashed line represents the line of no detection power, equivalent to the proportion of the positive class (average precision = 0.02). **b** Density histograms of predicted probabilities for positive cases (upward) and negative cases (downward) of GOSE > 5 prediction at hospital discharge. *n* represents the number of unique observations pertaining to each case and the range of predicted probabilities is fixed on a narrow, near-zero range to demonstrate the low predicted probabilities of the model.

**Supplementary Table S5**: Count distributions of GOSE scores at 12 months post discharge per observation window.

| Obs. Window (hr) | Unique patients (*n*) | Total obs. (*n*) | Glasgow Outcome Scale – Extended (GOSE) at 12 months post hospital discharge | | | | | | | |
|---|---|---|---|---|---|---|---|---|---|---|
| | | | 1 | 2 | 3 | 4 | 5 | 6 | 7 | 8 |
| 0.05 | 63 | 649 | 273 (44.25%) | 15 (2.43%) | 119 (19.29%) | 102 (16.53%) | 36 (5.83%) | 0 | 48 (7.78%) | 24 (3.89%) |
| 0.1 | 63 | 648 | 272 (44.16%) | 15 (2.44%) | 119 (19.32%) | 102 (16.56%) | 36 (5.84%) | 0 | 48 (7.79%) | 24 (3.90%) |
| 0.15 | 63 | 647 | 272 (44.23%) | 15 (2.44%) | 119 (19.35%) | 101 (16.42%) | 36 (5.85%) | 0 | 48 (7.80%) | 24 (3.90%) |
| 0.2 | 63 | 645 | 272 (44.37%) | 15 (2.45%) | 118 (19.25%) | 101 (16.48%) | 36 (5.87%) | 0 | 48 (7.83%) | 23 (3.75%) |
| 0.25 | 63 | 644 | 271 (44.28%) | 15 (2.45%) | 118 (19.28%) | 101 (16.50%) | 36 (5.88%) | 0 | 48 (7.84%) | 23 (3.76%) |
| 0.3 | 63 | 642 | 271 (44.43%) | 15 (2.46%) | 117 (19.18%) | 101 (16.56%) | 36 (5.90%) | 0 | 48 (7.87%) | 22 (3.61%) |
| 0.35 | 63 | 642 | 271 (44.43%) | 15 (2.46%) | 117 (19.18%) | 101 (16.56%) | 36 (5.90%) | 0 | 48 (7.87%) | 22 (3.61%) |
| 0.4 | 63 | 641 | 270 (44.33%) | 15 (2.46%) | 117 (19.21%) | 101 (16.58%) | 36 (5.91%) | 0 | 48 (7.88%) | 22 (3.61%) |
| 0.45 | 63 | 640 | 269 (44.24%) | 15 (2.47%) | 117 (19.24%) | 101 (16.61%) | 36 (5.92%) | 0 | 48 (7.89%) | 22 (3.62%) |
| 0.5 | 63 | 636 | 267 (44.13%) | 15 (2.48%) | 116 (19.17%) | 101 (16.69%) | 36 (5.95%) | 0 | 48 (7.93%) | 22 (3.64%) |
| 1 | 63 | 624 | 262 (44.18%) | 15 (2.53%) | 113 (19.06%) | 98 (16.53%) | 36 (6.07%) | 0 | 47 (7.93%) | 22 (3.71%) |
| 2 | 63 | 599 | 249 (43.76%) | 14 (2.46%) | 110 (19.33%) | 94 (16.52%) | 35 (6.15%) | 0 | 45 (7.91%) | 22 (3.87%) |
| 3 | 63 | 564 | 238 (44.40%) | 13 (2.43%) | 103 (19.22%) | 86 (16.04%) | 33 (6.16%) | 0 | 43 (8.02%) | 20 (3.73%) |
| 6 | 63 | 488 | 202 (43.53%) | 12 (2.59%) | 91 (19.61%) | 73 (15.73%) | 30 (6.47%) | 0 | 38 (8.19%) | 18 (3.88%) |
| 9 | 62 | 405 | 171 (44.30%) | 11 (2.85%) | 73 (18.91%) | 58 (15.03%) | 26 (6.74%) | 0 | 33 (8.55%) | 14 (3.63%) |
| 12 | 60 | 334 | 140 (43.89%) | 9 (2.82%) | 64 (20.06%) | 44 (13.79%) | 22 (6.90%) | 0 | 28 (8.78%) | 12 (3.76%) |
| 15 | 59 | 259 | 106 (42.74%) | 8 (3.23%) | 54 (21.77%) | 30 (12.10%) | 18 (7.26%) | 0 | 24 (9.68%) | 8 (3.23%) |
| 18 | 58 | 190 | 72 (39.78%) | 7 (3.87%) | 43 (23.76%) | 20 (11.05%) | 14 (7.73%) | 0 | 19 (10.50%) | 6 (3.31%) |
| 24 | 15 | 62 | 20 (32.79%) | 5 (8.20%) | 19 (31.15%) | 0 | 6 (9.84%) | 0 | 11 (18.03%) | 0 |

### Supplementary Figure S5

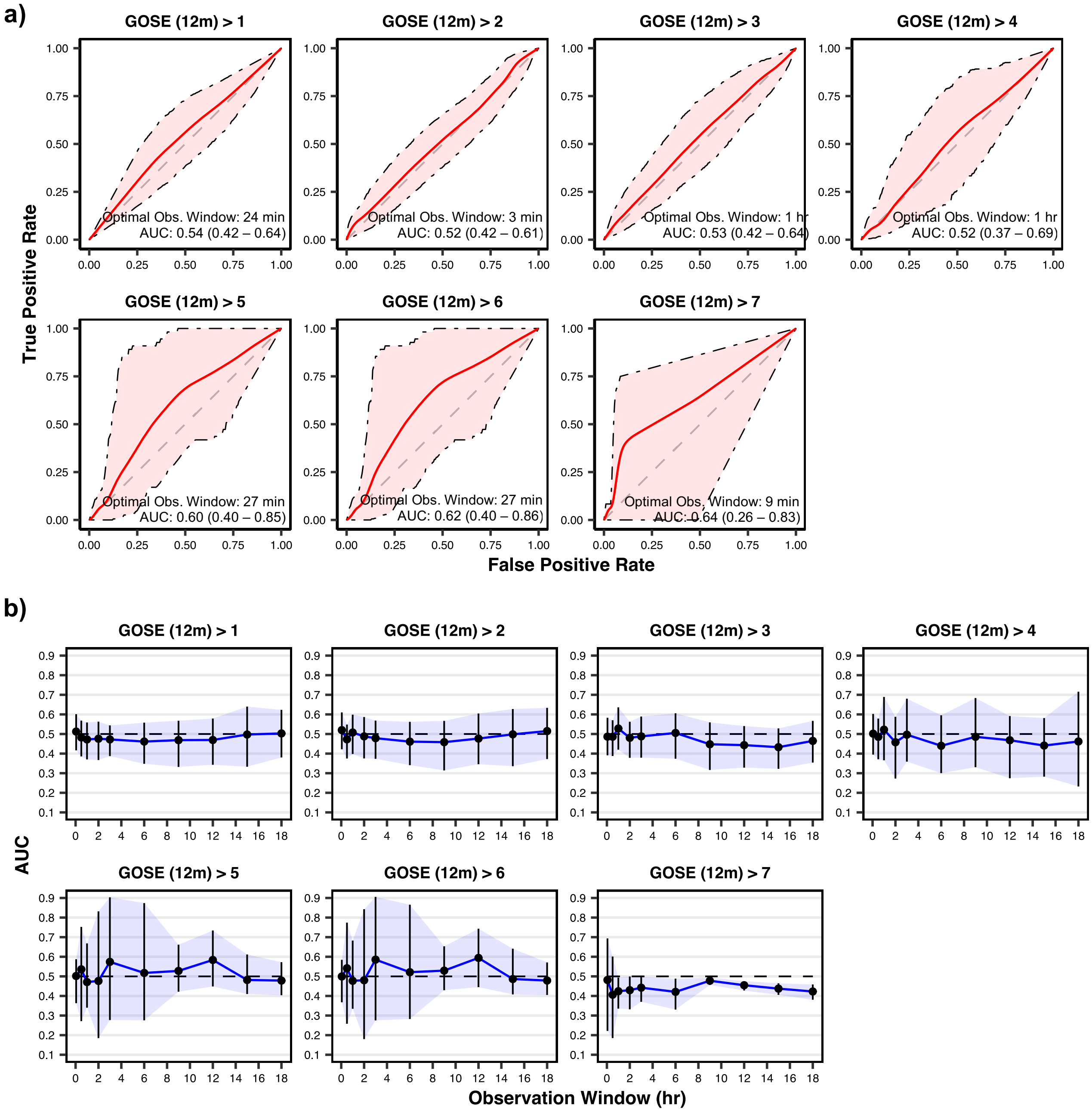


**Fig. S5. Discrimination performance of functional outcome at 12 months post discharge prediction models on validation sets. a** Receiver operating characteristic (ROC) curves of models pertaining to the observation windows with the highest achieved area under the ROC curve (AUC) per each tested prediction threshold of the Glasgow Outcome Scale – Extended (GOSE). Shaded areas represent 95% confidence intervals derived using bias-corrected bootstrapping (1,000 resamples) to represent the variation across repeated cross-validation folds (5 repeats of 5 folds) and nine missing value imputations. The values in each box represent the observation window achieving the highest AUC as well as the corresponding mean AUC (with 95% confidence interval in parentheses). The diagonal dashed line represents the line of no discrimination (AUC = 0.5). **b** AUC vs. observation windows up to 18 hours per each tested predicted threshold of the Glasgow Outcome Scale – Extended (GOSE). Points represent observation windows tested and error bars (with the associated shaded region) represent the 95% confidence interval. The horizontal dashed line corresponds to no discrimination (AUC = 0.5).

**Supplementary Table S6**: Discrimination of threshold-level GOSE at 12 months post discharge prediction models per observation window.

| Obs. Window (hr) | Unique patients (n) | Total observations (n) | AUC at different Glasgow Outcome Scale – Extended (GOSE) thresholds at 12 months post hospital discharge | | | | | | |
|---|---|---|---|---|---|---|---|---|---|
| | | | GOSE > 1 | GOSE > 2 | GOSE > 3 | GOSE > 4 | GOSE > 5 | GOSE > 6 | GOSE > 7 |
| 0.05 | 63 | 649 | 0.51 (0.42–0.60) | 0.52 (0.42–0.61) | 0.49 (0.39–0.58) | 0.50 (0.39–0.60) | 0.50 (0.36–0.59) | 0.50 (0.37–0.58) | 0.48 (0.22–0.69) |
| 0.1 | 63 | 648 | 0.51 (0.41–0.60) | 0.50 (0.40–0.59) | 0.50 (0.41–0.59) | 0.52 (0.39–0.63) | 0.49 (0.28–0.63) | 0.50 (0.30–0.64) | 0.53 (0.34–0.72) |
| 0.15 | 63 | 647 | 0.50 (0.41–0.57) | 0.47 (0.37–0.55) | 0.49 (0.40–0.58) | 0.51 (0.38–0.60) | 0.53 (0.36–0.69) | 0.54 (0.36–0.70) | 0.64 (0.26–0.83) |
| 0.2 | 63 | 645 | 0.50 (0.41–0.58) | 0.48 (0.38–0.57) | 0.48 (0.39–0.56) | 0.50 (0.36–0.69) | 0.49 (0.39–0.61) | 0.49 (0.39–0.63) | 0.49 (0.35–0.60) |
| 0.25 | 63 | 644 | 0.49 (0.40–0.57) | 0.51 (0.41–0.59) | 0.49 (0.39–0.59) | 0.49 (0.36–0.59) | 0.48 (0.39–0.59) | 0.49 (0.39–0.60) | 0.40 (0.21–0.50) |
| 0.3 | 63 | 642 | 0.50 (0.42–0.58) | 0.51 (0.40–0.59) | 0.49 (0.40–0.57) | 0.47 (0.36–0.56) | 0.55 (0.43–0.69) | 0.56 (0.43–0.70) | 0.51 (0.41–0.59) |
| 0.35 | 63 | 642 | 0.49 (0.39–0.58) | 0.50 (0.39–0.61) | 0.49 (0.39–0.59) | 0.49 (0.37–0.60) | 0.59 (0.42–0.80) | 0.58 (0.41–0.78) | 0.54 (0.40–0.67) |
| 0.4 | 63 | 641 | 0.54 (0.42–0.64) | 0.49 (0.40–0.58) | 0.48 (0.39–0.56) | 0.48 (0.33–0.62) | 0.59 (0.35–0.79) | 0.60 (0.35–0.80) | 0.46 (0.41–0.50) |
| 0.45 | 63 | 640 | 0.49 (0.38–0.57) | 0.50 (0.41–0.58) | 0.48 (0.38–0.58) | 0.47 (0.30–0.59) | 0.60 (0.40–0.85) | 0.62 (0.40–0.86) | 0.45 (0.38–0.50) |
| 0.5 | 63 | 636 | 0.48 (0.39–0.57) | 0.47 (0.38–0.55) | 0.49 (0.39–0.57) | 0.49 (0.37–0.58) | 0.54 (0.27–0.75) | 0.54 (0.26–0.77) | 0.41 (0.19–0.60) |
| 1 | 63 | 624 | 0.47 (0.37–0.56) | 0.51 (0.40–0.60) | 0.53 (0.42–0.64) | 0.52 (0.37–0.69) | 0.47 (0.34–0.67) | 0.48 (0.33–0.68) | 0.42 (0.34–0.49) |
| 2 | 63 | 599 | 0.48 (0.37–0.56) | 0.49 (0.38–0.59) | 0.48 (0.38–0.56) | 0.46 (0.27–0.59) | 0.48 (0.18–0.83) | 0.48 (0.18–0.84) | 0.43 (0.33–0.50) |
| 3 | 63 | 564 | 0.47 (0.39–0.54) | 0.48 (0.37–0.57) | 0.49 (0.38–0.59) | 0.50 (0.36–0.68) | 0.57 (0.28–0.90) | 0.59 (0.28–0.91) | 0.44 (0.37–0.50) |
| 6 | 63 | 488 | 0.46 (0.35–0.56) | 0.46 (0.34–0.56) | 0.51 (0.37–0.61) | 0.44 (0.30–0.60) | 0.52 (0.28–0.87) | 0.52 (0.28–0.87) | 0.42 (0.33–0.49) |
| 9 | 62 | 405 | 0.47 (0.33–0.57) | 0.46 (0.31–0.57) | 0.45 (0.32–0.56) | 0.49 (0.33–0.68) | 0.53 (0.42–0.66) | 0.53 (0.43–0.65) | 0.48 (0.46–0.49) |
| 12 | 60 | 334 | 0.47 (0.34–0.58) | 0.48 (0.35–0.60) | 0.44 (0.33–0.54) | 0.47 (0.27–0.59) | 0.58 (0.45–0.73) | 0.59 (0.45–0.74) | 0.45 (0.43–0.48) |
| 15 | 59 | 259 | 0.50 (0.33–0.64) | 0.50 (0.34–0.63) | 0.43 (0.32–0.53) | 0.44 (0.28–0.58) | 0.48 (0.41–0.61) | 0.49 (0.41–0.64) | 0.44 (0.41–0.46) |
| 18 | 58 | 190 | 0.50 (0.38–0.62) | 0.51 (0.37–0.63) | 0.46 (0.35–0.57) | 0.46 (0.23–0.72) | 0.48 (0.40–0.57) | 0.48 (0.41–0.57) | 0.42 (0.38–0.46) |
| 24 | 15 | 62 | 0.48 (0.15–0.86) | 0.49 (0.09–0.83) | 0.31 (0.08–0.66) | 0.31 (0.10–0.64) | 0.29 (0.06–0.58) | 0.29 (0.07–0.55) | |

Values in the seven rightmost columns represent mean validation set area under the receiver operating characteristic curve (AUC) values with associated 95% confidence intervals in parentheses. Confidence intervals were derived using bias-corrected bootstrapping (1,000 resamples) and represent the variation across repeated cross-validation folds (5 repeats of 5 folds) and nine missing value imputations. Missing values designate insufficient diversity in endpoint labels to evaluate models of that observation window and threshold combination.

### Supplementary Figure S6

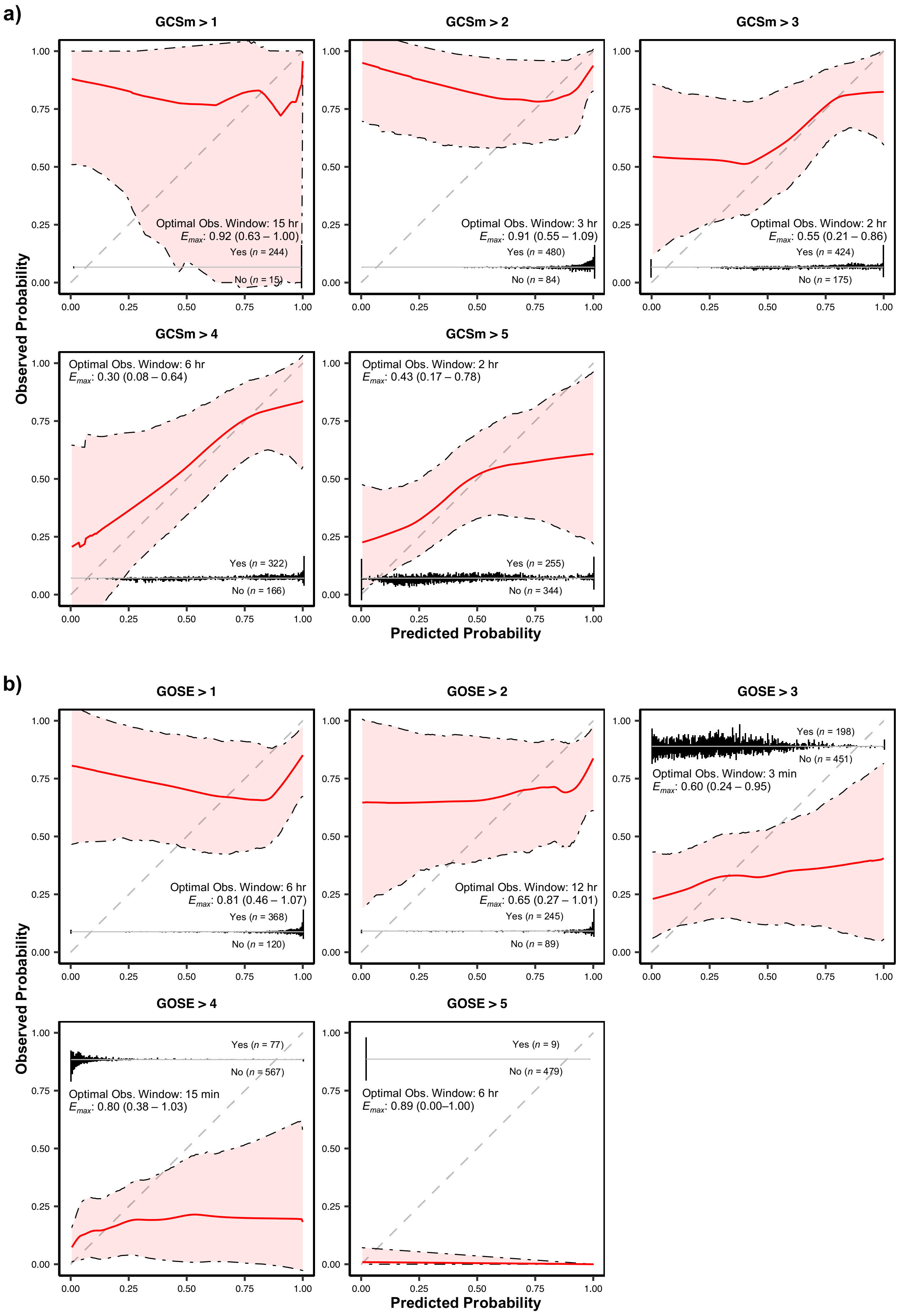


**Fig. S6. Probability calibration of optimally discriminating motor function detection and functional outcome prediction models on validation sets.** Validation set probability calibration curves of models pertaining to the observation windows with the highest achieved AUC (**a**) per each detection threshold of the motor component score of the Glasgow Coma Scale (GCSm) as shown in **Fig. 2a** and (**b**) per each tested prediction threshold of the Glasgow Outcome Scale – Extended (GOSE) as shown in **Fig. 3a**. Shaded areas represent 95% confidence intervals derived using bias-corrected bootstrapping (1,000 resamples) to represent the variation across repeated cross-validation folds (5 repeats of 5 folds) and nine missing value imputations. The distribution of predicted probabilities is shown at the bottom of the graphs, stratified by threshold-level endpoints. The values in each box represent the observation window achieving the highest AUC as well as the corresponding mean maximal absolute difference between observed and predicted probabilities of the endpoint ($E_{max}$) with 95% confidence interval in parentheses. The diagonal dashed line represents the line of perfect calibration.

**Supplementary Table 7**: Probability calibration metrics of optimally discriminating models.

| Task | Threshold | *n** | $E_{max}$ | $E_{50}$ | $E_{90}$ | ICI |
|---|---|---|---|---|---|---|
| Detection of GCSm | GCSm > 1 | 15/244 (0.94) | 0.92 (0.63–1.00) | 0.04 (0.01–0.10) | 0.04 (0.01–0.11) | 0.07 (0.02–0.12) |
| | GCSm > 2 | 84/480 (0.85) | 0.91 (0.55–1.09) | 0.09 (0.01–0.18) | 0.21 (0.09–0.36) | 0.11 (0.04–0.19) |
| | GCSm > 3 | 175/424 (0.71) | 0.55 (0.21–0.86) | 0.10 (0.02–0.21) | 0.25 (0.08–0.45) | 0.13 (0.05–0.22) |
| | GCSm > 4 | 166/322 (0.66) | 0.30 (0.08–0.64) | 0.10 (0.02–0.21) | 0.22 (0.06–0.44) | 0.11 (0.03–0.22) |
| | GCSm > 5 | 344/255 (0.57) | 0.43 (0.17–0.78) | 0.10 (0.03–0.23) | 0.31 (0.11–0.55) | 0.14 (0.06–0.24) |
| Prediction of GOSE at hospital discharge | GOSE > 1 | 120/368 (0.75) | 0.81 (0.46–1.07) | 0.17 (0.04–0.32) | 0.38 (0.19–0.61) | 0.20 (0.11–0.32) |
| | GOSE > 2 | 89/245 (0.73) | 0.65 (0.27–1.01) | 0.18 (0.04–0.34) | 0.39 (0.18–0.66) | 0.20 (0.10–0.32) |
| | GOSE > 3 | 451/198 (0.69) | 0.60 (0.24–0.95) | 0.14 (0.06–0.23) | 0.29 (0.14–0.45) | 0.15 (0.08–0.23) |
| | GOSE > 4 | 567/77 (0.88) | 0.80 (0.38–1.03) | 0.07 (0.02–0.17) | 0.15 (0.06–0.30) | 0.10 (0.05–0.18) |
| | GOSE > 5 | 479/9 (0.98) | 0.89 (0.00–1.00) | 0.00 (0.00–0.01) | 0.00 (0.00–0.01) | 0.01 (0.00–0.02) |
| Prediction of GOSE at 12 months post discharge | GOSE > 1 | 270/339 (0.56) | 0.63 (0.39–0.86) | 0.21 (0.11–0.32) | 0.49 (0.30–0.68) | 0.24 (0.15–0.34) |
| | GOSE > 2 | 288/329 (0.53) | 0.66 (0.43–0.88) | 0.19 (0.10–0.30) | 0.44 (0.29–0.59) | 0.22 (0.14–0.31) |
| | GOSE > 3 | 390/203 (0.66) | 0.66 (0.44–0.88) | 0.24 (0.11–0.38) | 0.57 (0.35–0.82) | 0.27 (0.19–0.35) |
| | GOSE > 4 | 488/105 (0.82) | 0.96 (0.76–1.06) | 0.14 (0.02–0.28) | 0.27 (0.10–0.47) | 0.17 (0.08–0.28) |
| | GOSE > 5 | 538/70 (0.88) | 0.87 (0.56–1.03) | 0.09 (0.01–0.20) | 0.22 (0.09–0.38) | 0.12 (0.04–0.24) |
| | GOSE > 6 | 538/70 (0.88) | 0.86 (0.53–1.03) | 0.09 (0.01–0.21) | 0.22 (0.08–0.36) | 0.13 (0.04–0.24) |
| | GOSE > 7 | 591/24 (0.96) | 0.92 (0.82–1.00) | 0.04 (0.01–0.07) | 0.04 (0.01–0.07) | 0.05 (0.02–0.09) |

Probability calibration metrics [mean (95% confidence interval)] corresponding to models trained on observation windows that maximize the area under the receiver operating characteristic curve (AUC) for each threshold (**Fig. 2a, 3a, and Supplementary Fig. S4 online**). Confidence intervals were derived using bias-corrected bootstrapping (1,000 resamples) and represent the variation across repeated cross-validation folds (5 repeats of 5 folds) and nine missing value imputations. Acronyms: motor component score of the Glasgow Coma Scale (GCSm), Glasgow Outcome Scale – Extended (GOSE), and Integrated Calibration Index (ICI).
*Count distribution of negative vs. positive cases with the proportion of the most represented case, equivalent to the no information rate, in parentheses.

### Supplementary Figure S7

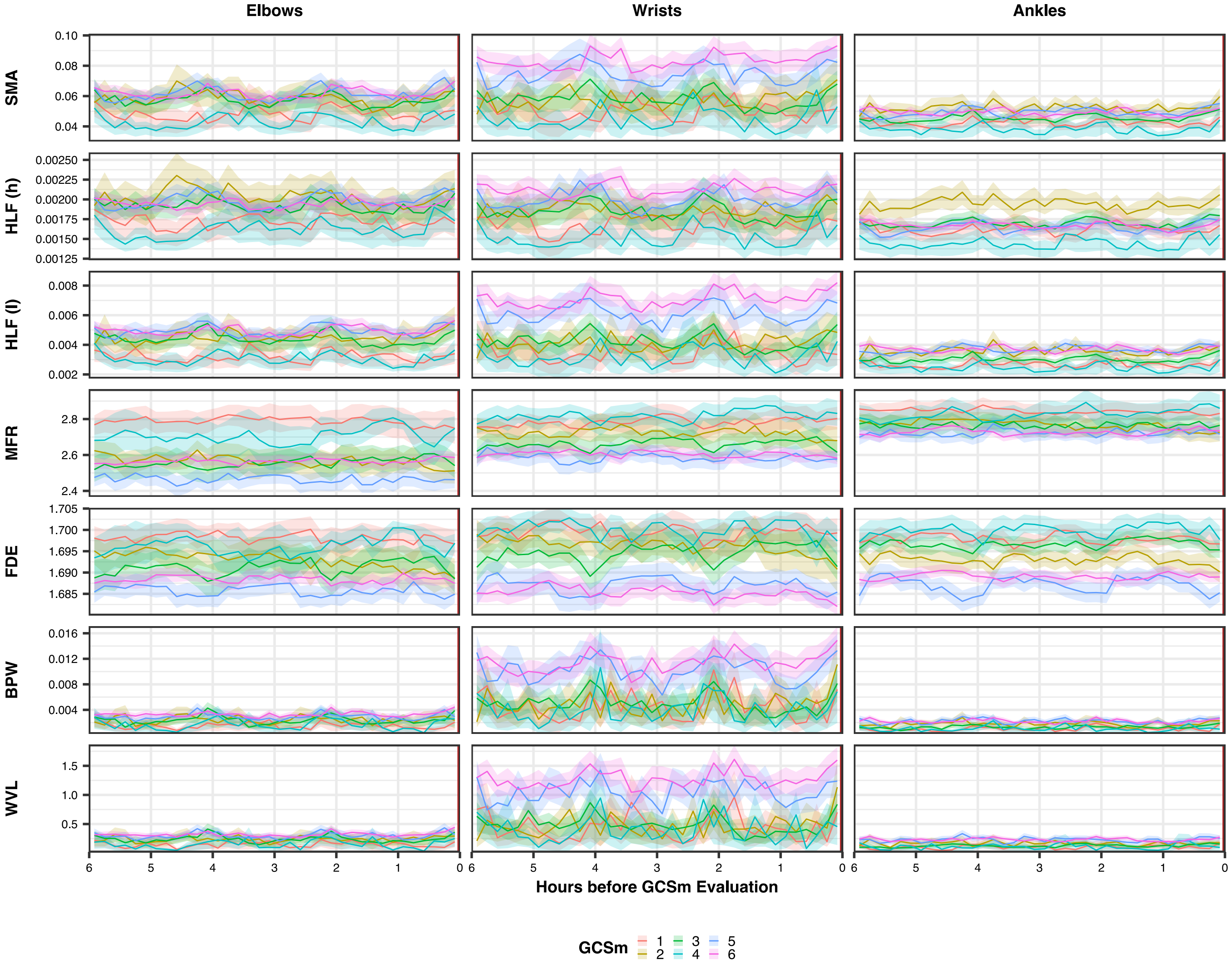


**Fig. S7. Mean motion feature trajectories in the six hours preceding GCSm evaluation, stratified by GCSm scores and bilateral sensor placement.** The figure represents the trajectories of features from 488 6-hour observation windows across 68 unique patients. Features have been binned in uniform 10-minute intervals preceding GCSm evaluation and outliers (values extending beyond three times the interquartile range above the third quartile of each combination of feature type and bilateral placement) were removed prior to calculation of the mean values. Shaded areas represent the 95% confidence interval bootstrapped from 1,000 resamples to represent the variation across unique GCSm observations. The solid dark red line on the rightmost edge of each graph represents the time of GCSm evaluation. Feature type acronyms are decoded in **Table 3**.

### Supplementary Figure S8

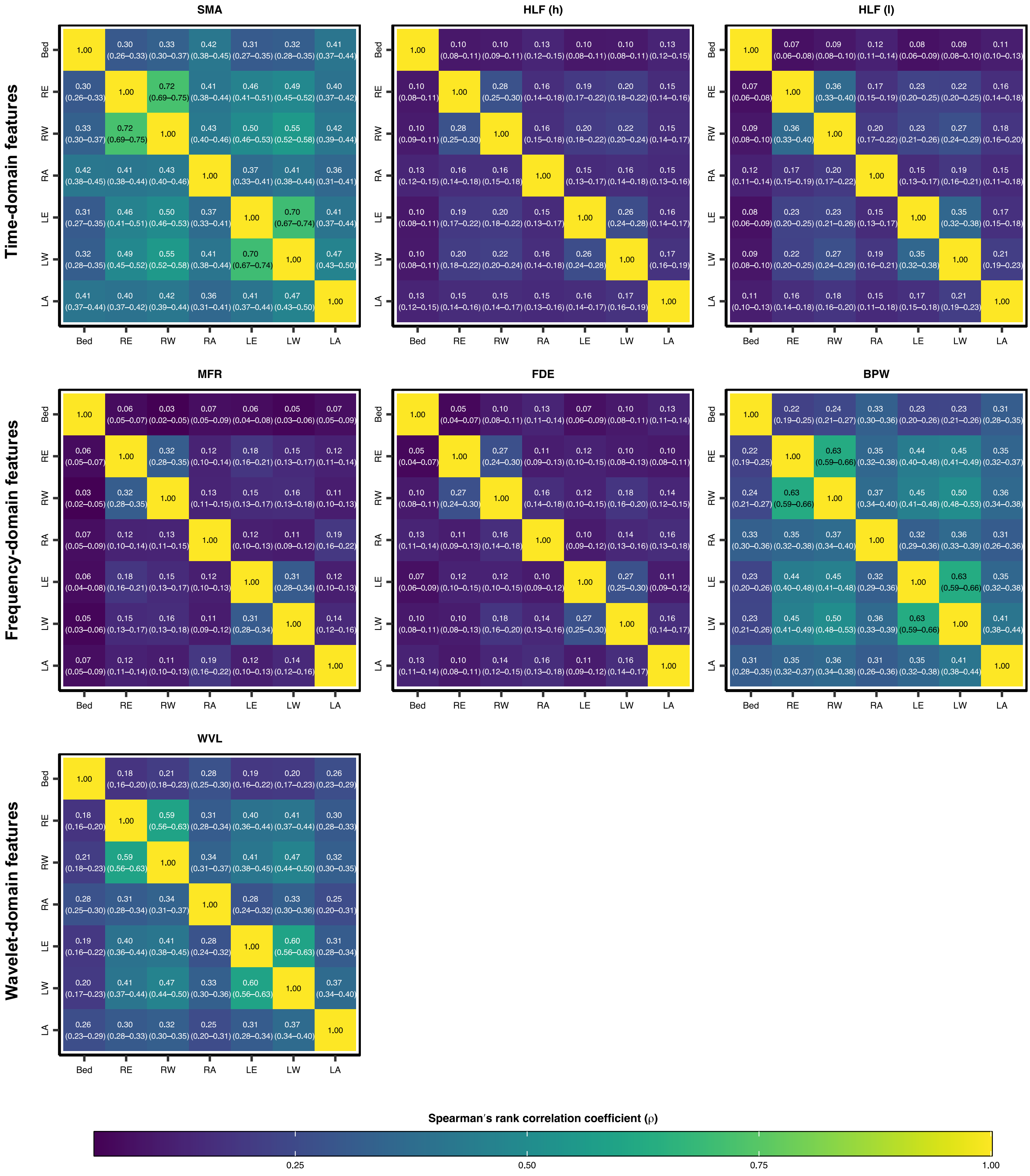


**Fig. S8. Correlation matrices of extracted motion features across different sensor placements.** Each matrix represents a unique feature type and values in each cell of the matrices represent the mean Spearman's rank correlation coefficient (ρ) between two sensor placements across the study population (*n* = 69) with the associated 95% confidence interval (bootstrapped with 10,000 resamples) in parentheses. Sensor placement acronyms correspond to the right and left elbows (RE and LE), the right and left wrists (RW and LW), and the right and left ankles (RA and LE). Feature type acronyms are decoded in **Table 3**.

### Supplementary Figure S9

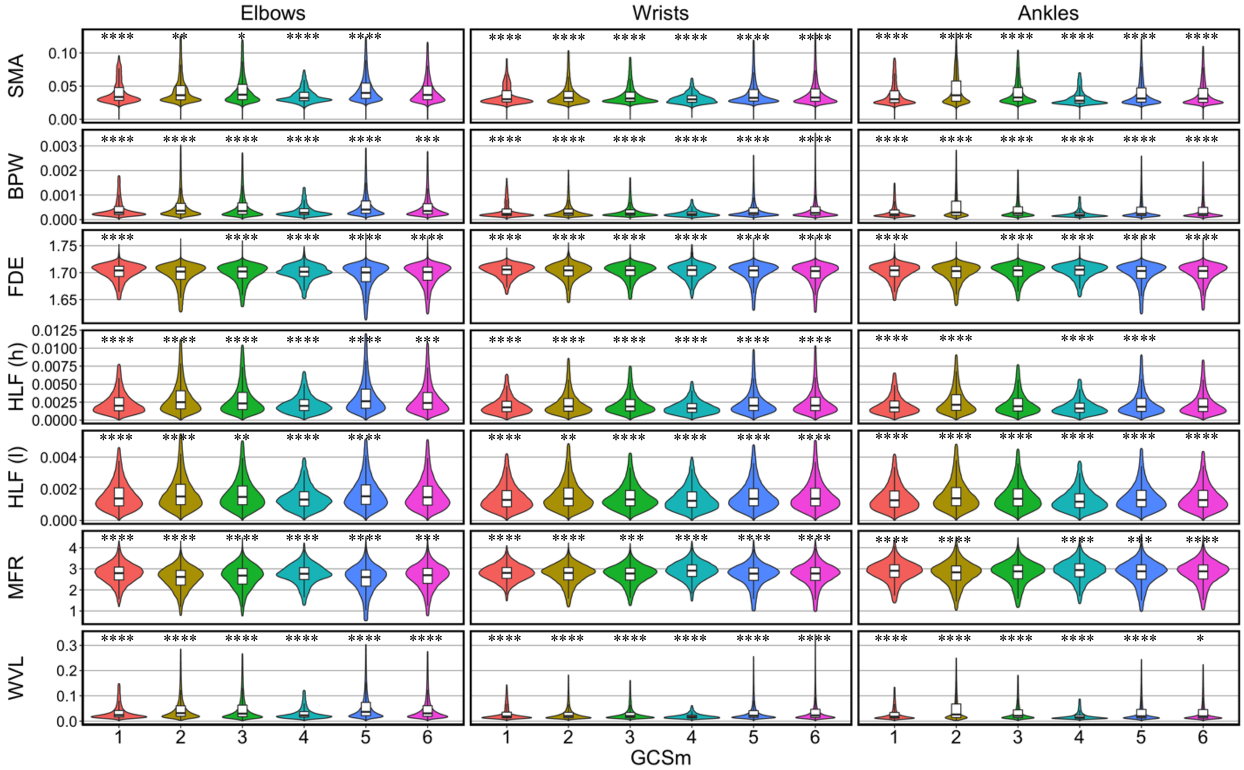


**Fig. S9. Violin plots of extracted motion feature values (30 min observation window), stratified by bilateral sensor placement and GCSm scores.** The figure represents the distribution of features from 636 observation windows across 68 unique patients. Outliers, defined as values extending beyond two times the interquartile range above the third quartile, were removed from the plot. Means of numerical distributions per GCSm score were each compared against the compiled distribution mean of all GCSm scores using the Wilcoxon signed-rank test. Statistically significant differences are marked with asterisks (*$p \leq 0.05$, **$p \leq 0.01$, ***$p \leq 0.001$, ****$p \leq 0.0001$). Feature type acronyms are decoded in **Table 3**.

**Supplementary Figure S10**

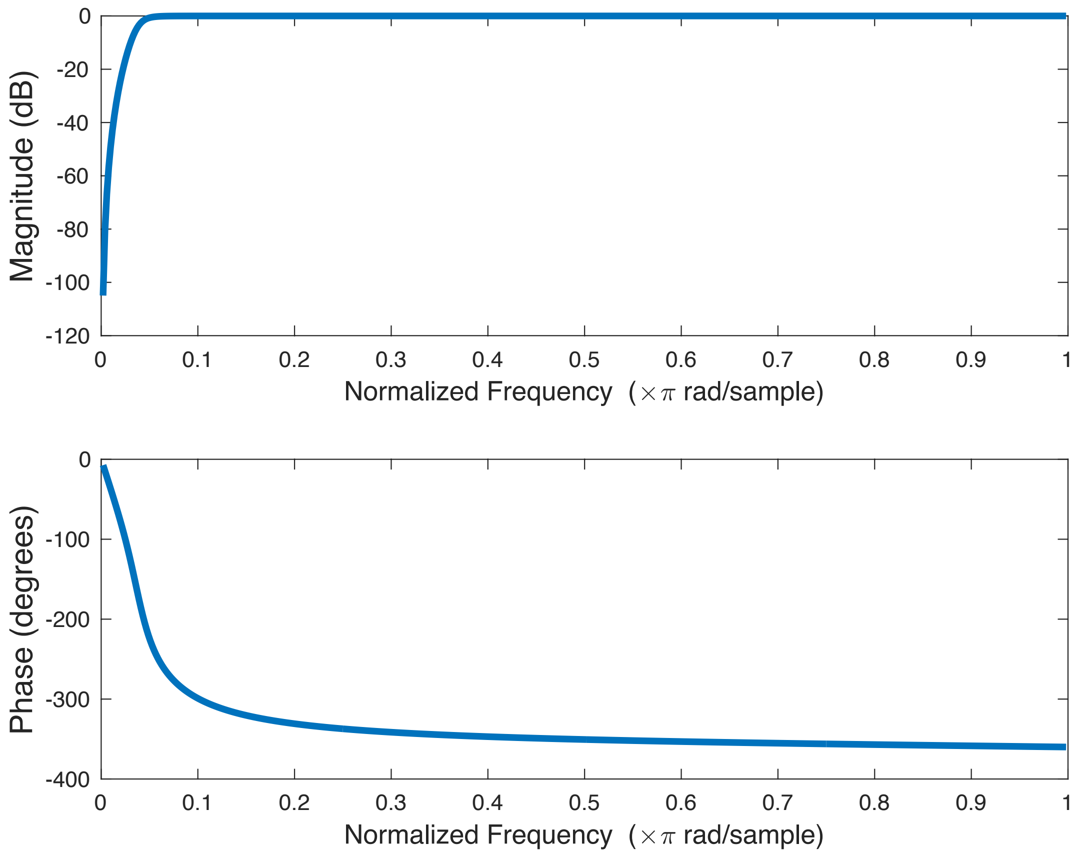


**Fig. S10. Bode plot of filter used in accelerometry processing.** *(top)* Magnitude response and *(bottom)* phase response of the 4th order Butterworth high-pass filter ($f_c$ = 0.2 Hz) used to filter out baseline offsets and static orientation from raw accelerometry ($f_s$ = 10 Hz).

**Supplementary Figure S11**

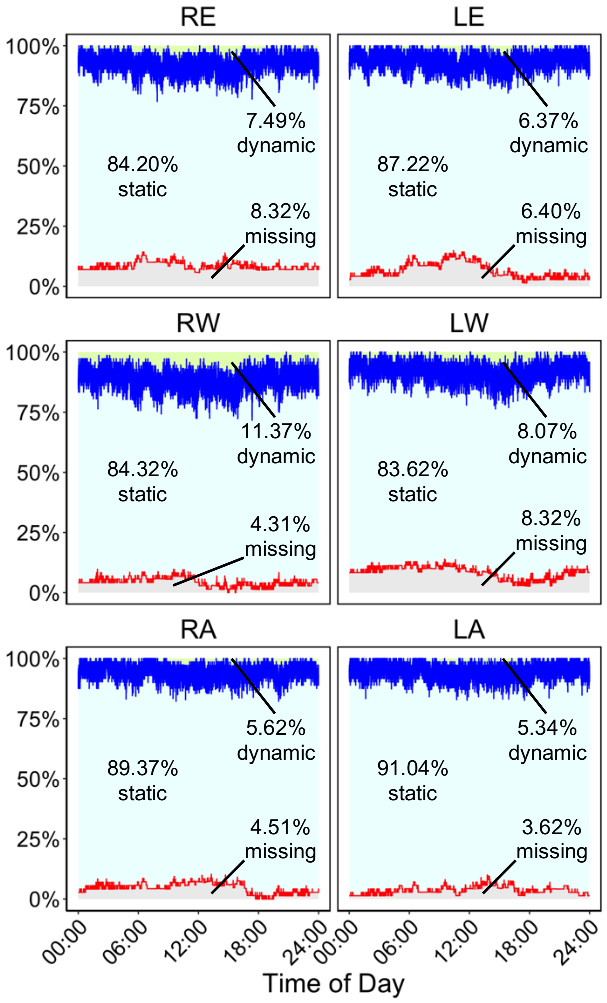


**Fig. S11. Percentages of missing, static, and dynamic accelerometry data by time of day of recording and sensor placement.** The complete accelerometry information (1,701 hours) across all patients ($n$ = 69) in the study were used to create this figure. The red line represents the percentage of missing data per time of day and the blue line represents percentage of missing data plus the percentage of static activity (SMA < 0.135 g) per time of day. Thus, the light grey shaded area represents the percentage of total missing data, the light cyan shaded area represents the percentage of total static activity, and the light green shaded area (barely visible) represents the percentage of total dynamic activity.

**Supplementary Table S8**: Percentages of missing accelerometry data per sensor and recording duration of each study participant.

| Patient Index | Bed | LA | LE | LW | RA | RE | RW | Recording Duration |
|---|---|---|---|---|---|---|---|---|
| 1 | 2.40% | 2.38% | 2.38% | 2.38% | 2.40% | 2.38% | 2.38% | 10:07:55 |
| 2 | 0.00% | 0.00% | 0.00% | 0.00% | 0.00% | 0.02% | 0.00% | 23:12:55 |
| 3 | 0.01% | 0.01% | 0.00% | 0.00% | 0.01% | 0.01% | 0.01% | 23:29:55 |
| 4 | 0.00% | 0.00% | 0.02% | 0.01% | 0.00% | 0.03% | 0.02% | 22:58:55 |
| 5 | 42.52% | 100.00% | 42.59% | 42.59% | 42.54% | 100.00% | 42.59% | 5:23:55 |
| 6 | 41.77% | 0.00% | 39.18% | 0.00% | 0.00% | 0.00% | 4.22% | 29:20:55 |
| 7 | 0.58% | 0.58% | 0.58% | 0.58% | 0.58% | 0.58% | 0.58% | 23:19:55 |
| 8 | 0.00% | 5.24% | 0.00% | 0.01% | 0.01% | 0.13% | 0.01% | 23:55:55 |
| 9 | 0.69% | 0.70% | 0.84% | 0.71% | 0.70% | 70.71% | 0.71% | 17:07:55 |
| 10 | 0.48% | 0.48% | 0.48% | 0.48% | 0.48% | 0.48% | 0.48% | 24:29:55 |
| 11 | 1.01% | 1.02% | 1.03% | 1.02% | 1.02% | 1.04% | 1.07% | 22:54:55 |
| 12 | 0.00% | 0.07% | 0.01% | 0.01% | 0.05% | 5.71% | 0.00% | 23:55:55 |
| 13 | 0.03% | 0.03% | 0.08% | 0.07% | 0.03% | 0.04% | 0.03% | 25:24:55 |
| 14 | 0.00% | 0.00% | 0.01% | 99.99% | 0.00% | 0.00% | 0.04% | 23:22:55 |
| 15 | 0.00% | 0.00% | 18.07% | 0.00% | 0.00% | 0.20% | 0.00% | 24:30:55 |
| 16 | 0.00% | 0.00% | 0.10% | 0.09% | 13.09% | 0.02% | 0.56% | 24:15:55 |
| 17 | 8.66% | 8.66% | 8.67% | 8.67% | 8.66% | 8.67% | 8.67% | 11:04:55 |
| 18 | 0.00% | 0.00% | 0.09% | 0.39% | 0.00% | 0.01% | 0.32% | 24:05:55 |
| 19 | 0.00% | 0.00% | 0.03% | 0.01% | 0.00% | 0.16% | 0.01% | 24:29:55 |
| 20 | 0.05% | 0.03% | 13.12% | 73.71% | 0.38% | 0.04% | 0.06% | 24:54:55 |
| 21 | 0.00% | 0.00% | 0.00% | 0.03% | 0.00% | 0.00% | 0.00% | 21:57:55 |
| 22 | 1.30% | 1.30% | 1.30% | 76.57% | 1.33% | 42.14% | 2.73% | 22:56:55 |
| 23 | 15.52% | 15.11% | 15.11% | 15.11% | 15.11% | 15.11% | 15.11% | 11:37:55 |
| 24 | 2.62% | 0.00% | 55.60% | 56.89% | 0.00% | 100.00% | 0.00% | 42:43:55 |
| 25 | 0.25% | 0.25% | 0.25% | 0.25% | 0.25% | 0.25% | 0.25% | 35:49:55 |
| 26 | 0.00% | 10.74% | 0.00% | 0.00% | 0.00% | 87.93% | 0.00% | 29:09:55 |
| 27 | 2.77% | 2.80% | 2.80% | 68.99% | 2.80% | 2.81% | 84.29% | 20:39:55 |
| 28 | 1.10% | 1.13% | 1.14% | 1.14% | 70.77% | 45.73% | 1.13% | 26:34:55 |
| 29 | 0.00% | 0.00% | 0.02% | 0.03% | 0.00% | 75.53% | 15.41% | 24:26:55 |
| 30 | 0.00% | 0.00% | 0.00% | 0.01% | 0.01% | 0.01% | 0.01% | 25:56:55 |
| 31 | 0.00% | 0.01% | 0.00% | 0.02% | 0.00% | 0.00% | 0.00% | 22:59:55 |
| 32 | 1.45% | 1.47% | 1.49% | 1.53% | 69.49% | 6.63% | 1.47% | 25:20:55 |
| 33 | 2.68% | 2.71% | 2.74% | 2.70% | 2.71% | 2.73% | 56.57% | 23:45:55 |
| 34 | 100.00% | 14.98% | 0.96% | 0.99% | 0.93% | 0.94% | 0.94% | 45:04:55 |
| 35 | 100.00% | 0.00% | 0.00% | 0.19% | 0.16% | 0.09% | 0.12% | 24:53:55 |
| 36 | 0.00% | 0.10% | 0.02% | 0.01% | 0.00% | 0.00% | 0.00% | 28:40:55 |
| 37 | 79.97% | 0.04% | 0.97% | 0.89% | 1.63% | 0.89% | 0.87% | 25:40:55 |
| 38 | 1.23% | 1.24% | 1.26% | 1.26% | 1.25% | 3.09% | 1.26% | 22:18:55 |
| 39 | 0.00% | 0.00% | 0.00% | 0.00% | 0.03% | 0.00% | 0.00% | 22:41:55 |
| 40 | 0.00% | 0.00% | 0.00% | 0.00% | 0.00% | 0.00% | 0.04% | 23:58:55 |
| 41 | 0.00% | 0.00% | 0.04% | 0.06% | 2.65% | 0.09% | 0.04% | 47:22:55 |
| 42 | 0.84% | 0.88% | 0.86% | 0.88% | 0.88% | 0.88% | 0.88% | 21:52:55 |
| 43 | 1.33% | 1.35% | 1.37% | 1.34% | 1.35% | 1.40% | 1.35% | 25:19:55 |
| 44 | 4.38% | 4.39% | 4.39% | 4.39% | 4.38% | 4.39% | 4.39% | 10:18:55 |
| 45 | 0.00% | 21.60% | 0.00% | 0.04% | 0.00% | 0.02% | 0.00% | 24:36:55 |
| 46 | 0.00% | 7.85% | 52.22% | 0.00% | 0.00% | 0.00% | 100.00% | 22:22:55 |
| 47 | 0.00% | 0.00% | 0.00% | 0.00% | 9.70% | 0.00% | 0.00% | 20:58:55 |
| 48 | 0.00% | 0.01% | 0.03% | 0.01% | 0.12% | 0.01% | 0.00% | 21:17:55 |
| 49 | 0.00% | 0.00% | 0.00% | 0.00% | 60.20% | 0.09% | 0.07% | 24:29:55 |
| 50 | 6.89% | 6.89% | 6.89% | 6.89% | 6.89% | 6.89% | 6.89% | 23:31:55 |
| 51 | 5.47% | 0.02% | 29.83% | 100.00% | 0.01% | 0.02% | 0.05% | 22:17:55 |
| 52 | 1.63% | 0.00% | 0.05% | 0.01% | 0.00% | 0.02% | 0.03% | 24:06:55 |
| 53 | 0.00% | 0.00% | 0.01% | 0.04% | 0.08% | 0.02% | 0.04% | 23:15:55 |
| 54 | 0.00% | 0.00% | 0.00% | 0.00% | 0.00% | 0.01% | 0.04% | 23:07:55 |
| 55 | 0.06% | 0.06% | 0.06% | 0.06% | 0.06% | 0.06% | 0.06% | 25:09:55 |
| 56 | 0.00% | 0.00% | 0.00% | 0.10% | 0.00% | 0.24% | 0.26% | 20:31:55 |
| 57 | 5.73% | 5.60% | 0.86% | 5.85% | 5.65% | 5.67% | 5.64% | 25:09:55 |
| 58 | 0.30% | 0.30% | 0.30% | 0.30% | 0.30% | 0.30% | 0.30% | 36:11:55 |
| 59 | 99.99% | 3.62% | 0.71% | 0.75% | 0.79% | 0.78% | 0.75% | 43:49:55 |
| 60 | 0.00% | 47.70% | 40.15% | 0.30% | 0.00% | 0.01% | 0.05% | 44:18:55 |
| 61 | 2.46% | 2.52% | 2.54% | 14.15% | 2.53% | 2.55% | 2.57% | 26:05:55 |
| 62 | 99.99% | 1.36% | 38.75% | 1.38% | 1.36% | 1.48% | 1.46% | 20:46:55 |
| 63 | 0.00% | 0.07% | 1.42% | 0.20% | 0.00% | 0.09% | 0.09% | 22:44:55 |
| 64 | 0.00% | 15.18% | 0.05% | 0.00% | 0.00% | 0.10% | 0.08% | 24:02:55 |
| 65 | 72.66% | 0.01% | 0.00% | 0.01% | 0.00% | 0.03% | 0.01% | 23:55:55 |
| 66 | 0.00% | 0.05% | 0.00% | 0.00% | 0.00% | 0.00% | 0.05% | 22:53:55 |
| 67 | 0.00% | 0.00% | 34.26% | 0.29% | 0.01% | 0.00% | 0.09% | 23:15:55 |
| 68 | 0.09% | 0.00% | 0.00% | 15.78% | 0.00% | 0.00% | 2.74% | 23:55:55 |
| 69 | 1.03% | 1.04% | 1.03% | 1.05% | 20.98% | 1.04% | 1.11% | 23:11:55 |
| *Total* | 11.82% | 3.62% | 6.40% | 8.32% | 4.51% | 8.32% | 4.31% | 1701:00:15 |

Recording duration is specified in hours:minutes:seconds. The total percentage of missing data, across all patients and all sensors, is 6.76% (excluding bed sensor: 5.91%).

**Supplementary Table S9**: Ranges of static activity values for each motion feature.

| Feature | Minimum Value | Maximum Value |
|---|---|---|
| SMA | 0.000 | 0.135 |
| $HLF_h$ | 0.000 | 0.008 |
| $HLF_l$ | 0.000 | 0.006 |
| MFR | 1.630 | 3.200 |
| FDE | 1.630 | 1.710 |
| BPW | 0.000 | 0.012 |
| WVL | 0.000 | 1.000 |

The maximum value (0.135) of SMA was proposed by Lugade et al. For the remaining feature spaces, we determined the threshold of dynamic activity by minimizing the Euclidean norm of the proportion of static activity across the patient set with that of the SMA threshold. Feature type acronyms are decoded in **Table 3**.
